# Pragmatic modeling supports current dosing guidelines for carbamazepine and valproic acid for the treatment of epilepsy in children

**DOI:** 10.1101/2024.12.13.24318984

**Authors:** Joyce E.M. van der Heijden, Violette Gijsen, Anne M. van Uden, Marika de Hoop-Sommen, Jolien J.M. Freriksen, Elke Jacobs, Rick Greupink, Saskia N. de Wildt

**Affiliations:** Department of Pharmacy - Pharmacology Toxicology, Radboud university medical center, Nijmegen, The Netherlands; HagaZiekenhuis-Juliana Childrens Hospital; Department of Neonatal and Pediatric Intensive Care, Division of Pediatric Intensive care, Erasmus MC-Sophia Children’s Hospital, Rotterdam, The Netherlands; Department of Intensive Care, Radboud university medical center, Nijmegen, The Netherlands

**Keywords:** Carbamazepine, Valproic acid, Epilepsy, Modeling & simulation, Pediatrics, Pharmacokinetics, Physiologically-based pharmacokinetic modeling

## Abstract

**Background:** Carbamazepine (CBZ) and valproic acid (VPA) are long-standing treatments for epilepsy in children. Interestingly, they display unique drug disposition characteristics and maturation of drug metabolizing enzymes further complicates personalized dosing. Physiologically-based pharmacokinetic (PBPK) modeling includes these mechanisms and is hence a promising tool to optimize dosing. Our aim is to better support pediatric drug dosing of CBZ and VPA.

**Methods:** All CBZ and VPA dosing simulations were conducted with Simcyp, using available CBZ and VPA compound models linked with adult and pediatric population models. Current Dutch national dosing strategies were simulated to evaluate their appropriateness to achieve therapeutic levels. Where doses could be optimized, alternative dosing strategies were proposed based on simulations.

**Results:** Therapeutic levels of CBZ and VPA will be reached after 1 or 2 weeks of treatment with the current dosing strategies. Simulations suggest a CBZ starting dose of 7 mg/kg/day for neonates rather than 10 mg/kg/day. In contrast, children aged 12 to 18 years may receive a higher starting dose (e.g., 400 mg/day instead of 200 mg/day), to reach therapeutic levels more quickly. For VPA, when higher doses are needed (i.e., ≥30 mg/kg/day), measuring unbound VPA concentrations are advised to guide dosing.

**Conclusion:** We demonstrate that PBPK modeling is a valuable tool to confirm and further optimize dosing recommendations in children. The use of PBPK modeling offers a practical, cost-effective, and swift method to provide valuable comprehensive evidence for guiding clinical practice and potentially informing pediatric drug labeling, thus eliminating the necessity for clinical studies.

## Introduction

Epilepsy remains a prevalent neurological disorder in children, affecting 0.5-1% of children during their childhood (1). Adequate antiepileptic pharmacotherapy can ensure maximal seizure control, with the ultimate goal of seizure freedom, while mitigating the risk of adverse effects and maintaining a good quality of life. The choice of antiepileptic drug (AED) will be determined by factors such as the type of epilepsy, the child’s age, side-effects, sex, and the potential for drug-drug interactions (DDIs). For generalized seizures, valproic acid (VPA) is often prescribed, while for focal seizures, carbamazepine (CBZ) is internationally commonly chosen (2, 3). Both drugs are first generation AEDs that have been prescribed in children for decades. Current dosing regimens for both CBZ and VPA follow a step-up dosing protocol in which each individual is clinically evaluated to determine when the desired effect is observed (4–7).

The dosing of these drugs can be quite complex, as they possess unique drug disposition mechanisms that may be even more complex in the pediatric population. Drug absorption, clearance, and the formation of active metabolites, such as carbamazepine-10,11-epoxide (CBZE) and 4-ene-valproic acid (4-ene-VPA), can either assist or hinder effective seizure control (8–10). VPA is extensively bound to albumin in a concentration-dependent manner and is metabolized by multiple cytochrome P450 (CYP) and uridine 5’-diphospho-glucuronosyltransferase (UGT) enzymes (8, 11). In contrast, CBZ is mainly metabolized by CYP3A4 and induces its own metabolism (auto-induction) (9). These characteristics result in a high interindividual variation in CBZ and VPA levels.

In addition, age-related physiological changes can significantly impact pharmacokinetics (PK) and thereby influence drug and metabolite concentrations and thereby efficacy and toxicity (12). For example, age-related changes in drug metabolizing enzyme activity or changes in renal function warrant age-appropriate doses to ensure optimal drug efficacy and safety. By incorporating knowledge of age-related variations in drug disposition processes into PK models, age-appropriate dosing recommendations can be established (13). Pediatric physiologically-based pharmacokinetic (PBPK) models account for these age-related physiological changes and improve continuously with increasing knowledge (14). PBPK modeling is increasingly accepted to guide dosing in pediatric clinical care and is also supported by regulatory agencies, such as the US Food and Drug Administration and European Medicines Agency to underpin first-in-child dosing during pediatric drug development (15–17).

Moreover, PBPK modeling facilitates the exploration of numerous ‘what if’ scenarios that would otherwise necessitate extensive, costly, logistically challenging, and sometimes ethically questionable clinical studies. As indicated, VPA is highly albumin-bound, and albumin levels may vary between subjects. A population PK study has shown that albumin levels have a significant impact on VPA clearance and, therefore, albumin levels should be checked before dosing (18). Moreover, several case studies including patients with hypoalbuminemia show VPA toxicity despite therapeutic total VPA levels (19, 20). Exploration with PBPK modeling may enable quantitative characterization of these ‘what-if’ scenarios, informing improved VPA dosing recommendations.

Given their routine use in clinical practice and the availability of abundant pediatric clinical data, CBZ and VPA serve as ideal proof-of-concept drugs to demonstrate the value of the PBPK modeling approach in pediatric neurology to provide additional insights to obtain model-informed doses or to strengthen currently recommended doses. We have previously demonstrated that a pragmatic PBPK modeling approach is feasible and have described the workflow in detail (21, 22). Our aim is to provide more insight into pediatric drug dosing of CBZ and VPA, including altered albumin levels on VPA exposures.

## Methods

### Step 1. Adult and pediatric model verification

#### Model selection

To conduct PBPK simulations, we used Simcyp PBPK software v21 for CBZ and v22 for VPA (Certara UK Limited, Simcyp Division, Sheffield, UK), a population-based PBPK modeling platform. The software already contains a well-validated adult population model and a pediatric population model with age-related varying physiology, including relevant CYP and UGT enzyme ontogeny profiles (i.e., CYP2C8, CYP2C9, CYP3A4, UGT1A4, UGT1A6, UGT1A9, and UGT2B7). These population models were linked to 1) a CBZ immediate release (IR) compound model with the CBZE metabolite (Table S1), 2) a VPA IR compound model (23), and 3) a VPA extended release (ER) compound model (24), with both VPA models including the 4-ene-VPA metabolite (Table S2).

#### Model verification

To verify that these models adequately predict CBZ and VPA concentrations in adults and subsequently in children across the pediatric age span, we first searched PubMed for adult and pediatric PK data to compare predicted CBZ and VPA concentrations with observed data (Table S3-10). The accuracy of model predictions (i.e., predictive performance) was assessed quantitatively by calculating predicted-to-observed PK parameter ratios (i.e., C_max_, C_min_, C_av_, T_max_, AUC, Vd, CL, t_1/2_; within 2-fold range was considered acceptable) and qualitatively by a visual predictive check comparing the predicted and the observed plasma concentration-time curves. A more detailed description of the model verification process can be found in the supplementary materials.

### Step 2. Dose simulations

After model verification, the models were applied prospectively to evaluate current dosing strategies for oral administration in children. CBZ and VPA dosages for epilepsy, as provided in the Dutch Paediatric Formulary (DPF), were simulated (4, 5). For VPA, only one bodyweight-based dosing recommendation is available for a broad age range (i.e., 1 month – 18 years); therefore, separate simulations were conducted with smaller age ranges to explore the age-related effects on predicted drug exposure in more detail. The simulated age ranges include 0-1 month, 1 month to 1 year, 1-5 years, 5-12 years, and either 12-16 years and 16-18 years (for CBZ), or 12-18 years (for VPA), which are based on the current CBZ dose recommendations in the DPF.

The therapeutic targets chosen for our simulations were for CBZ and VPA a trough concentration (the plasma concentration immediately before the next dose is administered; C_trough_) of 4-12 μg/ml and 50-100 μg/ml, respectively (25). For CBZE, the upper boundary of the reference range, i.e., 2.3 μg/ml, is included (26). Based on reference concentrations of 50-100 μg/ml total VPA, reference values for the free concentration are 4-15 μg/ml (i.e., 7-9% unbound at 50 μg/ml and 15% at 100 μg/ml total concentrations) (11, 27). Determination of the total plasma concentration is sufficient for VPA in most cases, although, it may occasionally be of relevance to determine the free fraction, such as in cases of unexpected toxic effects with total VPA concentrations in the therapeutic reference range or hypoalbuminemia (28). Although 4-ene-VPA is associated with VPA therapy-related hepatotoxicity (29), no clear reference threshold is defined. One study suggests a minimum toxic plasma concentration of above 0.5 μg/ml for 4-ene-VPA (30). Nevertheless, both CBZE and 4-ene-VPA metabolite thresholds are included in the evaluation.

### Step 3. Effect of variation in albumin levels on total and unbound VPA exposure

To explore the effect of altered albumin levels in children on VPA PK, the mean total VPA, unbound VPA, and 4-ene-VPA concentrations were simulated under conditions of +20%, average,-20%, and-35% age normalized reference albumin levels. Additionally, the effects of different doses (i.e., 7.5, 10, 12.5, and 15 mg/kg q12h IR) in the context of different albumin levels were explored.

## Results

To determine whether the PBPK models could accurately predict CBZ, CBZE, total VPA (IR and ER), unbound VPA, and 4-ene-VPA in adults and children, predictions were compared with observed data. Figure 1 provides mean predicted plasma concentration-time profiles in children upon multiple oral administrations of CBZ, IR VPA, and ER VPA, all indicating accurate predictions. Furthermore, the majority of predicted-to-observed PK parameter ratios falling within the 2-fold range (Figure S2-5). To note, CBZ multi-dose simulations performed much better compared to single-dose simulations in both adults and children (Table S11). Additional adult and pediatric verification data are available in the supplementary materials (Figure S6-14).

**Figure 1.**
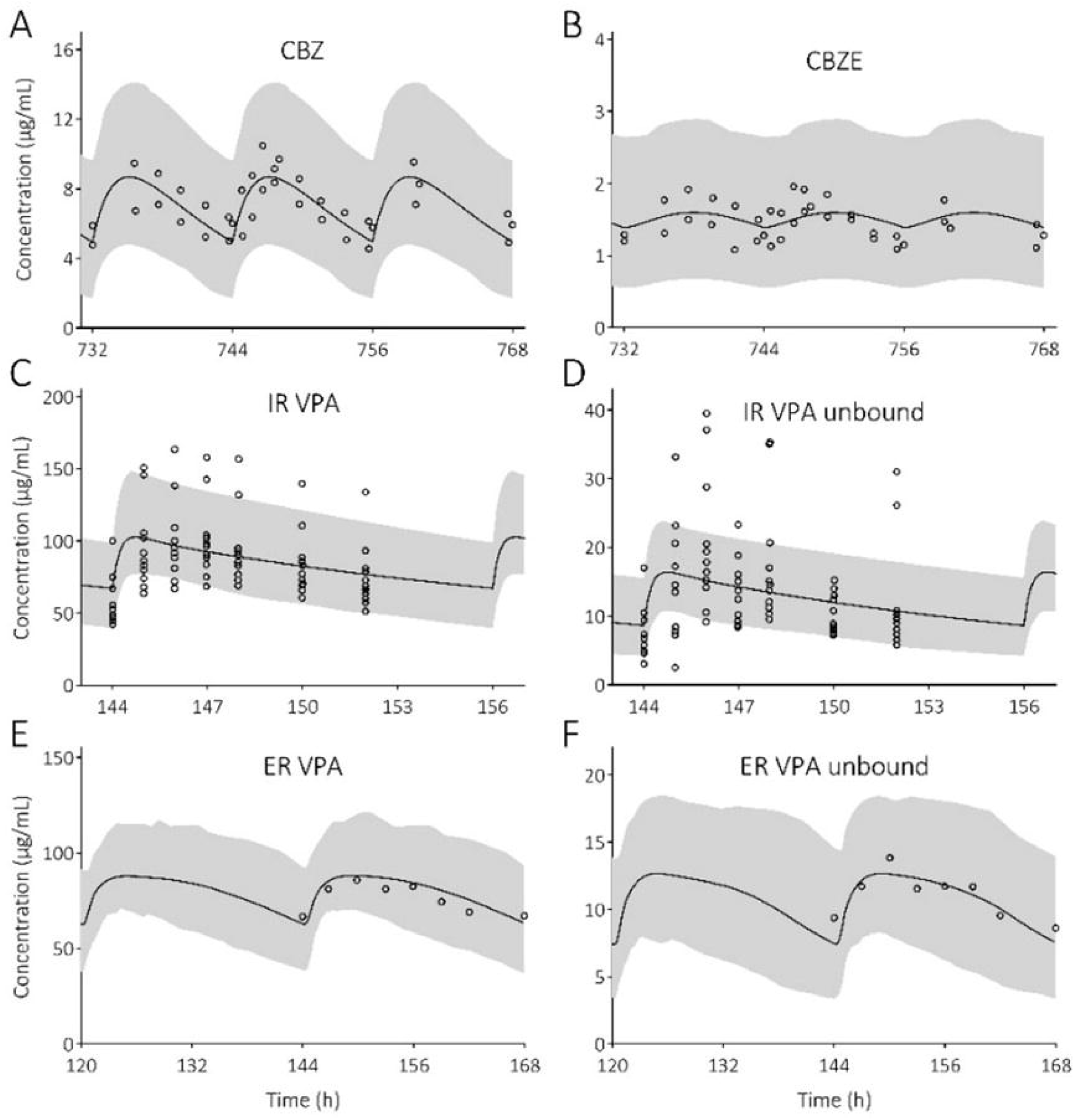
Visual predictive checks of predicted carbamazepine (CBZ), carbamazepine-10,11-epoxide (CBZE), valproic acid (VPA) and unbound VPA plasma concentration-time profiles compared to observed clinical data in pediatric patients. Solid line is the predicted mean and the shaded area represents the 5^th^-95^th^ percentile of the predicted plasma concentration in the virtual population. Open circles are the observed data after the following oral doses: A&B) 7.0 mg/kg every 12 hours (q12h) (∼7-16 years) (1), C&D) immediate-release (IR) 14.83 mg/kg q12h (0.5-1.5 years) (2), and E&F) extended-release (ER) 19.9 mg/kg every 24 hours (8-11 years) (3). A, B, C and D provide individual data points, whereas E and F are mean values.

After model verification, current dosing strategies were simulated and evaluated against the therapeutic targets and the upper boundary of the reference range for the metabolites. For CBZ, most children are expected to reach therapeutic C_trough_ levels after either 1 or 2 weeks of treatment (Figure 2). When higher doses (i.e., 25 mg/kg/day) appear clinically needed to achieve a beneficial effect in children aged 1 to 12 years, one should be cautious regarding the less favorable the CBZE/CBZ ratio (Figure 2). Furthermore, simulations showed that neonates should receive a starting dose of 7 mg/kg/day to prevent concentrations exceeding the upper range of the therapeutic target for a large proportion of neonates instead of the higher 10 mg/kg which is also used (Figure S17). From a PK point of view, children aged 12 to 18 years may receive a higher starting dose, for example 400 mg/day instead of 200 mg/day to reach therapeutic levels earlier (Figure S18).

**Figure 2.**
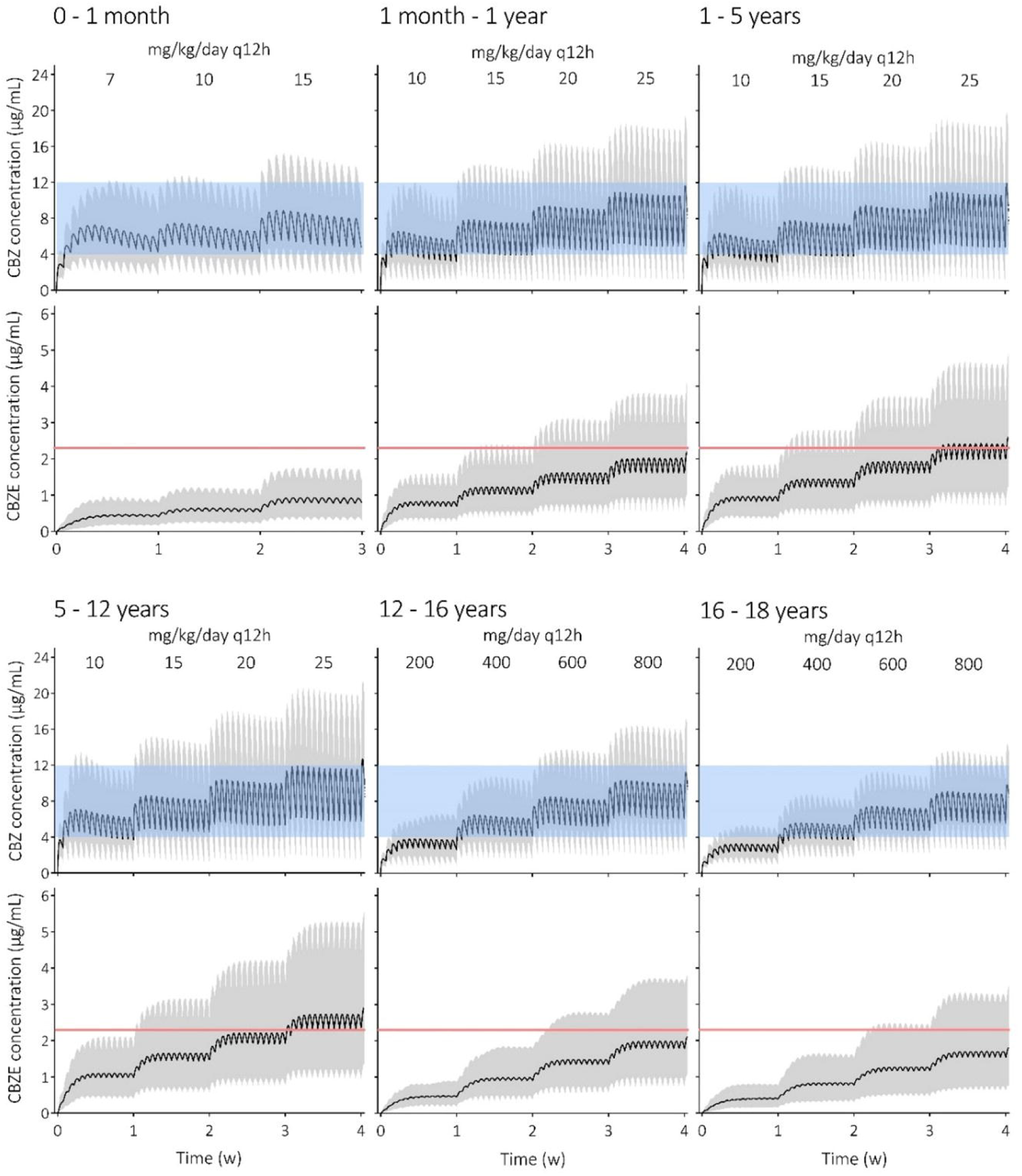
Prediction of carbamazepine (CBZ) and carbamazepine-10,11-epoxide (CBZE) concentrations following the current Dutch Paediatric Formulary dosing guidelines. Solid black line is the predicted mean and the shaded area represents the 5^th^-95^th^ percentile of the predicted plasma concentration in the virtual population. The blue shaded area indicates the CBZ therapeutic window (i.e., 4-12 μg/ml) and the red solid line indicates the CBZE upper boundary of the reference range (i.e., 2.3 μg/ml). Abbreviation: every 12 hours (q12h).

For VPA, approximately half of the virtual children (all age ranges) reached therapeutic C_trough_ levels after 1 week of treatment with 20 mg/kg/day (IR q12h or ER q24h; Figure 3). Unbound VPA levels (both upon IR and ER) and 4-ene-VPA levels (only upon IR) were also adequate after 1 or 2 weeks of treatment (Figure S19-23). When exploring total VPA and unbound VPA concentrations upon altered albumin levels per age group, mean total VPA concentrations dropped below the therapeutic target with reduced albumin levels (i.e.,-20 and-35%), whereas unbound levels remained within the therapeutic window (Figure S24). When adjusting the dose to improve total VPA concentrations with altered albumin levels (e.g., 15 mg/kg q12h under-35% albumin conditions), mean total VPA concentrations nicely fell within the therapeutic window, yet unbound VPA concentrations could still exceed the maximum therapeutic concentration due to an altered unbound fraction (Figure 4). The observed concentrations can be explained by the altered VPA (hepatic) clearances upon altered albumin levels (Figure S25) and by minor alterations in volume of distributions (Figure S26). Therefore, we advise to routinely determine unbound VPA concentrations in patients with hypoalbuminemia and/or higher VPA doses to monitor free VPA toxicity.

**Figure 3.**
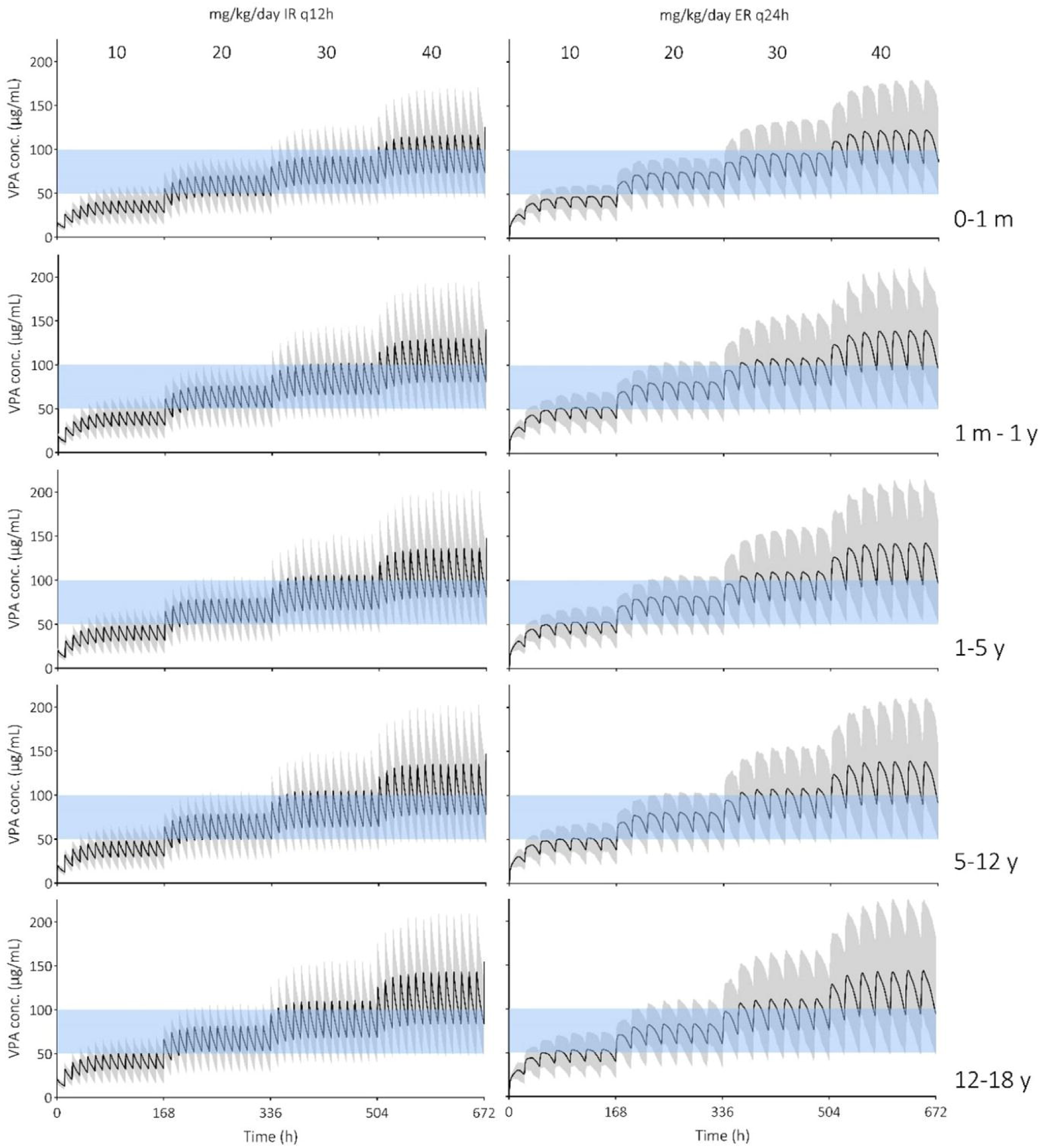
Prediction of valproic acid (VPA) concentrations following the current Dutch Paediatric Formulary immediate release (IR) and extended release (ER) dosing guidelines. Solid black line is the predicted mean and the shaded area represents the 5^th^-95^th^ percentile of the predicted plasma concentration in the virtual population. The blue shaded area indicates the VPA therapeutic window (i.e., 50-100 μg/ml). Abbreviations: month (m), year (y), every 12 hours (q12h), every 24 hours (q24h).

**Figure 4.**
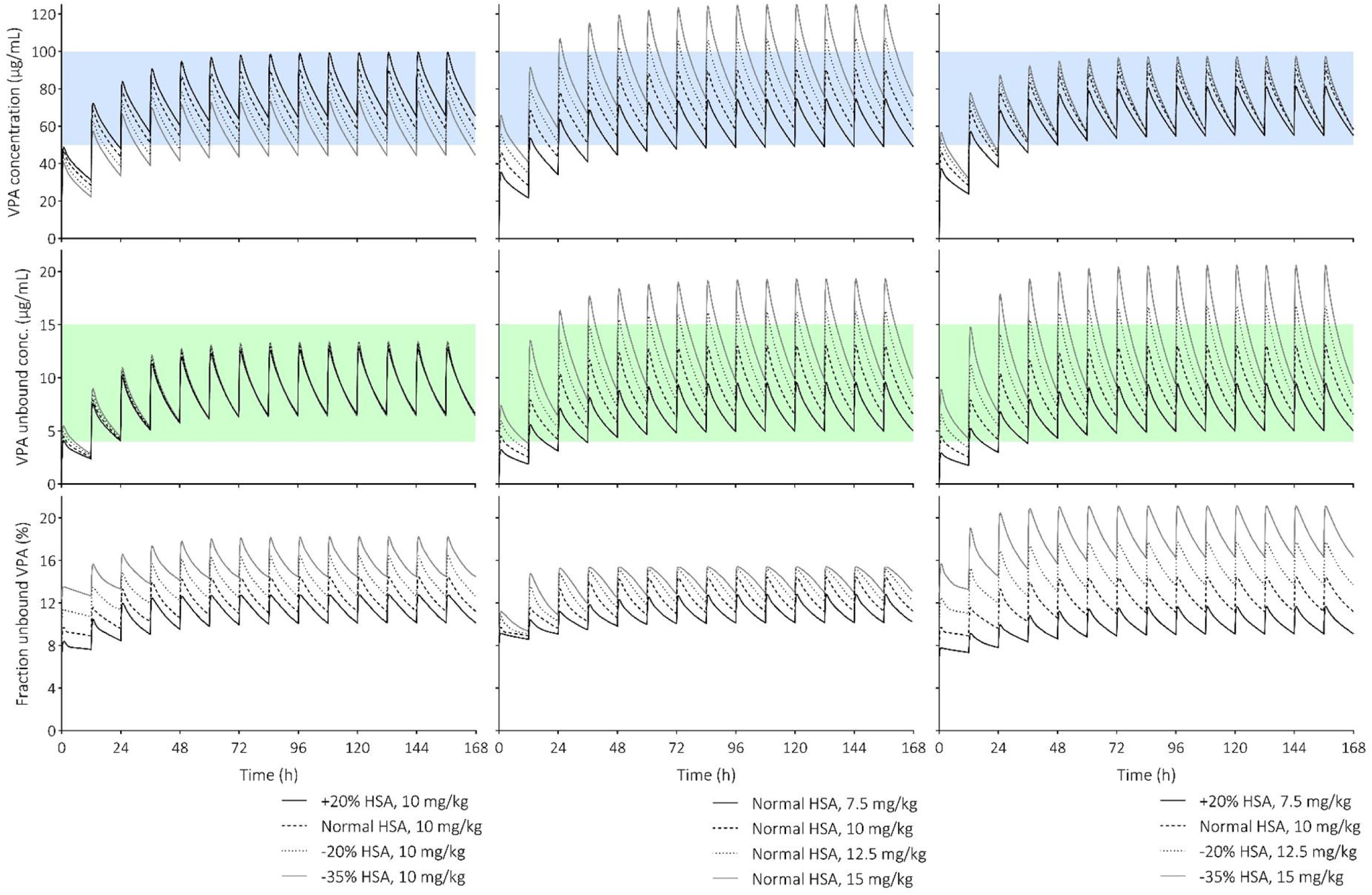
Prediction of valproic acid (VPA) and unbound VPA concentrations, as well as fraction unbound following varying immediate-release dosages and adjusted human serum albumin (HSA) levels in children 12-18 years. The lines are the predicted means of the simulated populations. The blue shaded area indicates the VPA therapeutic window (i.e., 50-100 μg/ml) and the green shaded area indicates the unbound VPA therapeutic window (i.e., 4-15 μg/ml).

## Discussion

In this study, we applied PBPK modeling and simulation to evaluate current DPF CBZ and VPA dosing recommendations in the treatment of epilepsy in children. By assessing predicted drug and primary metabolite concentrations against established therapeutic targets and toxicity thresholds, respectively, it was confirmed that current national dosing recommendations, including step-up dosing, appear adequate for reaching appropriate levels for children of all ages. As our simulations show, due to the large interindividual variability in drug exposure between children, increasing the dose until each individual achieves a clinically desirable effect remains the preferred approach. Some finetuning of the current dose recommendations can be advised and will be discussed in more detail below.

First, it is advised to initiate neonatal CBZ treatment with 7 mg/kg/day instead of 10 mg/kg/day. Other guidelines providing CBZ dosing recommendations for neonates are lacking, complicating the comparison of our suggested model-informed dose. In contrast, children aged 12 to 18 years may receive a higher starting dose (e.g., 400 mg/day instead of 200 mg/day). Our suggestion to increase initial CBZ doses in children aged 12-18 years compared to the current guidelines is logical from a PK viewpoint, aiming to achieve therapeutic concentrations earlier. Lexicomp suggests an initial CBZ dose of 200 mg twice daily in adolescents (31), this is in line with our suggestion to initiate treatment with 400 mg/day instead of 200 mg/day. Still, standard of care is to initiate treatment with a low dose to reduce side effects. This is frequently described in literature, yet underlying evidence is poor as it is still mostly anecdotal (32). Drowsiness, ataxia, and vertigo are commonly observed upon therapy initiation or upon dose increase (33). Yet, as the actual difference in incidences and administered dosages are not specified in the study, it is still unclear if a higher starting dose would lead to more side effects. As underlying data is ambiguous, we suggest that physicians should carefully consider each patient’s need for rapid seizure control against the anticipated side effects and, consequently, compliance to therapy.

Furthermore, additional attention is needed when higher doses are needed to achieve a beneficial effect, as the CBZE/CBZ ratio becomes less favorable. An inverse relationship between age and CBZE/CBZ ratio was observed in children (0-18 years), resulting in a higher CBZE/CBZ ratio compared to adults (34, 35), which aligns with our findings. Additionally, with higher CBZ doses, auto-induction of CYP3A4 increases, consequently leading to increased CBZE formation, while CBZE elimination remains constant, ultimately resulting in higher CBZE/CBZ ratios. Higher CBZE/CBZ ratios have been associated with a higher incidence of side effects (36, 37), but this could not be confirmed by other studies (38, 39). CBZE undergoes epoxide hydroxylation to the inactive trans-10,11-dihydrodiol CBZ (40). Little is known regarding the ontogeny of epoxide hydrolase. Yet what is known is that its expression in the fetal liver is approximately 25-50% of adult levels (41, 42), and hepatic enzymatic levels increase linearly with gestational age (43). However, further information on its development in children is unknown and it has been argued that the degradation of CBZE is age-independent (44). Within the model, CBZE elimination is not mechanistically incorporated as it is described as an oral clearance based on adult clinical data with a 30% coefficient of variance. The oral clearance follows allometric scaling (using an allometric exponent of 0.75 and a reference adult body weight of 70 kg), but whether this holds true for epoxide hydrolase activity is unknown. Despite this uncertainty, CBZE concentrations were predicted rather accurately with the pediatric model, exhibiting only a slight underprediction and a greater predicted variability compared to the observed data (Figure S9). Moreover, CBZE data were only available in children aged 4-17 years, and evaluated doses ranged from 6.75-10 mg/kg q12h (Table S6). Therefore, a level of uncertainty in CBZE model predictions is present in children younger than 4 years old and doses above 10 mg/kg q12h. Overall, we believe it is necessary to be cautious regarding elevated CBZE levels in younger children, especially following high CBZ doses.

For VPA, a dosing strategy of 20 or 30 mg/kg/day upon IR and ER administration seems to yield appropriate levels of total VPA, unbound VPA and 4-ene-VPA across all simulated age groups (Figure 3 & S19-23). Interestingly, predicted VPA levels are slightly lower in the 0-1 month age group compared to the other pediatric age groups receiving the same bodyweight-based dose, indicating a higher predicted clearance when corrected for bodyweight. Clearances from observed data are highest within the 1-2 year old age group, and decreases to adult levels at approximately 12 years of age (45, 46). Comparing the predicted clearance across the pediatric age span with observed clearance values shows that the increase in clearance within the 1-2 year age span is not adequately captured (Figure S27). This discrepancy was unnoticed during model verification as the included studies represented predominantly broad age ranges. Only one study including children aged 0.5-1.5 years (n=7) found a higher observed clearance compared to our predicted clearance values (i.e., predicted/observed ratios were 0.76-0.86 fold) (47). Additionally, predicted VPA concentrations accurately matched observed data (Figure 2). Although predictions are within acceptable ranges, the observed data indicate higher clearances or oral absorption in children aged 1-2 years and might therefore require higher doses.

A total VPA concentration within the therapeutic window does not necessarily indicate that the unbound (reflecting active drug) concentration is within its therapeutic window. Several case reports and case series have indicated that hypoalbuminemia causes VPA toxicity despite a therapeutic total VPA level (19, 20). Another study indicated that patients with elevated unbound VPA levels had significantly lower median serum albumin levels than those with unbound VPA levels within the therapeutic range (48). Our simulations indicated that altering the albumin levels only changes total VPA levels, while having no effect on unbound VPA levels, due to compensatory alteration in clearances and fraction unbound. Yet, in case the VPA dose is adjusted based on total VPA levels, it can indeed result in undesirable elevated unbound peak VPA levels, even within normal albumin ranges (i.e.,-20%) (Figure 4). Therefore, we advise to routinely determine unbound VPA concentrations in patients with hypoalbuminemia and/or higher VPA doses (i.e., above 30 mg/kg/day) to monitor free VPA toxicity.

Several population PK (popPK) models have previously been published for VPA. Three VPA popPK studies indicated that children aged 1-2 years require higher bodyweight-based VPA doses compared to older children. According to Ding et al., children aged 1 year should receive a VPA daily dose of approximately 20 mg/kg/day to reach a total VPA steady-state C_trough_ of 50 μg/ml (49), whereas Gu et al. recommended a 50 mg/kg/day dose to achieve therapeutic unbound VPA levels (almost 75% of the simulated population has a C_trough_ above 4 μg/ml) (50). Simulations from Tauzin et al. suggested a dose of at least 40 mg/kg/day for patients ≤10 kg (is equal to <1 year age), yet half of the simulated patients remained underdosed (51). In contrast, our simulations suggest that the majority of children aged 1 year reach therapeutic total and unbound VPA C_trough_ levels with 30 mg/kg/day (Figure S19-23). A possible cause why we do not observe such an age-related effect with our PBPK model, is that the fraction metabolized by an additional hepatic clearance input (non-mechanistic input value) is between 50-60% for children aged 0-5 years, potentially overshadowing the maturational effect of the included CYP and UGT ontogeny profiles in the model.

Our study has some limitations. Firstly, the therapeutic windows used for both CBZ and VPA are based on clinical experience. Since the relationship between plasma level and efficacy is unpredictable for both drugs (26), no therapeutic drug monitoring (TDM) is generally recommended (27, 52). Only in cases of unexplained side effects or lack of effect TDM is used, adhering to the therapeutic windows presented here. This does not alter the fact that in practice both lower and higher levels have been shown to be effective. Nonetheless, a PK target is required to aim for during dose finding simulations and, therefore, applied here as a directive.

Secondly, AEDs are often not administered as monotherapy and AED drug-drug interactions are frequent. Additionally, AEDs have more DDIs than drugs of other therapeutic classes, posing a major challenge in treatment (53). However, in this study, no DDI simulations have been performed, and recommendations are made only for CBZ and VPA monotherapy. The effect of concomitant use of other AEDs can be explored with pragmatic PBPK modeling, as PBPK compound models are available for certain AEDs that include enzyme kinetics and inhibition/induction data, such as lamotrigine (54) and phenytoin (Simcyp software).

Thirdly, an abundant number of papers was available for both CBZ and VPA model verification in children (i.e., 10 and 15 studies, respectively), yet certain age ranges are still lacking. We lack adequate PK data in children for CBZ single dose between 1-5 years, PK data for CBZ multi dose <4 years, and PK data for VPA ER <4 years of age. Conducting clinical studies to fill these information gaps, if feasible, may take years to conduct. While we rely on well validated PBPK models based on virtual physiology that is constantly refined over time and by delineating relevant PK mechanisms, are we able to provide pragmatic solutions. Extrapolating modeling predictions for the age groups lacking observed data was necessary. Yet, despite this, the reliability in model performance is high as also our predictions are in line with current clinical practice.

To conclude, we demonstrate that pragmatic PBPK modeling is a valuable tool for providing additional insights for model-informed doses for both CBZ and VPA in the treatment of epilepsy in children. Our data indicate that therapeutic levels of CBZ and VPA are expected to be achieved after 1 or 2 weeks of treatment with the current DPF dosing strategies. In addition, this study provides suggestions to optimize CBZ doses in neonates and children aged 12-18 years, along with the recommendation to assess unbound VPA levels if high VPA doses are required or in case of reduced albumin levels. The use of PBPK modeling and comparing predicted concentrations against established therapeutic windows offers a practical, cost-effective, and swift method to provide valuable comprehensive evidence for guiding clinical practice and potentially informing pediatric drug labeling, thus eliminating the necessity for clinical studies.

## Acknowledgements

We thank Dr. Ping Zhao for useful discussions that improved the article.

## Data Availability

The raw data supporting the conclusions of this article will be made available by the authors, without undue reservation.

