## Supplemental material for "Pragmatic modeling supports current dosing guidelines for carbamazepine and valproic acid for the treatment of epilepsy in children"

#### 1. Model input parameters

**Table 1.** Drug-dependent input parameters for carbamazepine (CBZ) and carbamazepine-10,11-epoxide (CBZE)

| | Parameter | CBZ | CBZE |
|---|---|---|---|
| Physicochemical properties | Molecular weight (g/mol) | 236.27 | 252.27 |
| | Neutral species octanol : buffer partition coefficient (Log $P_{O:W}$) | 2.22 | 1.44 |
| | Compound type | Neutral | Neutral |
| | Blood-to-plasma partition ratio (B/P) | 1.07 | 1.53 |
| | Fraction unbound (fu) | 0.25 | 0.48 |
| | Plasma binding protein | HSA | HSA |
| Absorption | Absorption model | First-order absorption | |
| | Fraction available from dosage form (fa) | 0.84 | 0.84 |
| | First order absorption rate constant (ka; 1/h) | 0.5 | 0.19 |
| | Lag time (h) | 0 | 0 |
| | Unbound fraction of drug in enterocytes ($fu_{gut}$) | 1 | 1 |
| | A nominal flow in gut model ($Q_{gut}$; L/h) | 14.419 (P) | |
| | Human jejunum effective permeability ($P_{eff,man;}$ $10^{-4}$cm/s) | 4.3 | 1.7787 (P) |
| | Permeability Assay; values | | PSA ($Å^2$); 58.9, HBD; 2 |
| Distribution | Distribution model | Minimal PBPK model | Minimal PBPK model |
| | Volume of distribution at steady state ($V_{SS}$; L/kg) | 0.78 (30% CV) | 0.78 (30% CV) |
| Elimination | Oral clearance ($CL_{PO}$) (L/h) | | 6.07 (36 % CV) |
| | CYP3A4 – 10,11-epoxidation (Vmax; Km) | 0.72; 180.2 | |
| | CYP3A5 – 10,11-epoxidation (Vmax; Km) | 1.44; 332.3 | |
| | CYP2C8 – 10,11-epoxidation (Vmax; Km) | 0.03; 741.74 | |
| | CYP3A4 – Additional Inducible ($CL_{int}$) | 0.0106 | |
| | UGT2B7 – N-glucuronidation (Vmax; Km; fumic) | 0.79; 126.9; 0.2 | |
| | rUGT Scalar (Liver; Intestine; Kidney) | 5.29; 0.63; 0.85 | |
| | Additional Liver CL (HLM) (µL/min/mg protein) | 0.255 | |
| | Typical renal clearance ($CL_R$) (L/h) | 0.0084 | 0.14 |
| Interaction | CYP3A4 - $Ind_{slope}$^; CV (%); $fu_{inc}$ | 0.16; 30; 1 | 0.14; 30; 1 |
| | CYP3A5 - $Ind_{slope}$^; CV (%); $fu_{inc}$ | 0.16; 30; 1 | 0.14; 30; 1 |

**Table 2.** Drug-dependent input parameters for valproic acid (VPA) immediate-release (IR), VPA extended release (ER), and 4-ene-VPA

| | Parameter | VPA IR (1) | 4-ene-VPA (1) | VPA ER (2) |
|---|---|---|---|---|
| Physicochemical properties | MW (g/mol) | 144.21 | 142.2 | 144.211 |
| | Log $P_{O:W}$ | 2.75 | 2.21 | 2.75 |
| | pKa | 4.8 | 5.06 | 4.8 |
| | Compound type | Monoprotic acid | Monoprotic acid | Monoprotic acid |
| | B/P | 0.55 | 0.55 | 0.55 |
| | Concentration dependent fu* | 0.07-0.15 (3) | 0.0567 (P) | 0.07-0.15 (3) |
| | Plasma binding protein | HSA | HSA | HSA |
| Absorption | Absorption model | First-order | | ADAM |
| | Fa | 1 | | |
| | ka (1/h) | VPA = 2 | | |
| | | NaVPA / DVS = 4.1 | | |
| | Lag time (h) | EC tablets = 2 | | |
| | $fu_{gut}$ | 1 | | |
| | $Q_{gut}$ (L/h) | 16.0 (P) | | |
| | $P_{eff,man}$ ($10^{-4}$cm/s) | 5.8304 (P) | | 3.84 (P) |
| | Permeability Assay; values | PSA (Å$^2$); 37.3, HBD; 1 | | Caco-2 (6.5:7.4); 22.6, reference; propranolol, Caco-2 (6.5:7.4); 21.5 |
| Distribution | Distribution model | Minimal | Minimal | Minimal |
| | $k_{in}$ (1/h), $k_{out}$ (1/h), $V_{sac}$ (L/kg) | 0.217, 0.866, 0.036 | | 0.217, 0.866, 0.036 |
| | $V_{SS}$ (L/kg) Adult | 0.10843 | 0.209 | 0.14 |
| | $V_{SS}$ (L/kg) Pediatric, Kp Scalar | 0.19001, 2.254 | | |
| Elimination | IV clearance ($CL_{IV}$) (L/h) | | 2.601 | 0.52 |
| | rUGT Scalar | 0.37 | | 0.23 |
| | UGT1A3 ($V_{max}$; $K_m$) | 600; 6400 | | 600; 6400 |
| | UGT1A4 ($V_{max}$; $K_m$) | 400; 3100 | | 400; 3100 |
| | UGT1A6 ($V_{max}$; $K_m$) | 700; 3200 | | 700; 3200 |
| | UGT1A8 ($V_{max}$; $K_m$) | 870; 5900 | | 870; 5900 |
| | UGT1A9 ($V_{max}$; $K_m$) | 960; 6300 | | 960; 6300 |
| | UGT1A10 ($V_{max}$; $K_m$) | 880; 5500 | | 880; 5500 |
| | UGT2B7 ($V_{max}$; $K_m$) | 1200; 1400 | | 1200; 1400 |
| | CYP2C9 – 4-ene-VPA ($CL_{int}$), fm | 0.0003976, 0.00785 | | 0.0015 |
| | CYP2C9 – Other ($CL_{int}$), fm | 0.0026609, 0.05255 | | |
| | Additional hepatic $CL_{int}$ | 0.2 | | 0.125 |
| | $CL_{add}$ (L/h) | | 0.3414 | |
| | $CL_R$ (L/h) | 0.0079 | 2.2596 | 0.0052 |

*Interpolation method: Piecewise Cubic Polynomial. Abbreviations: divalproex sodium (DVS), sodium valproate (NaVPA).

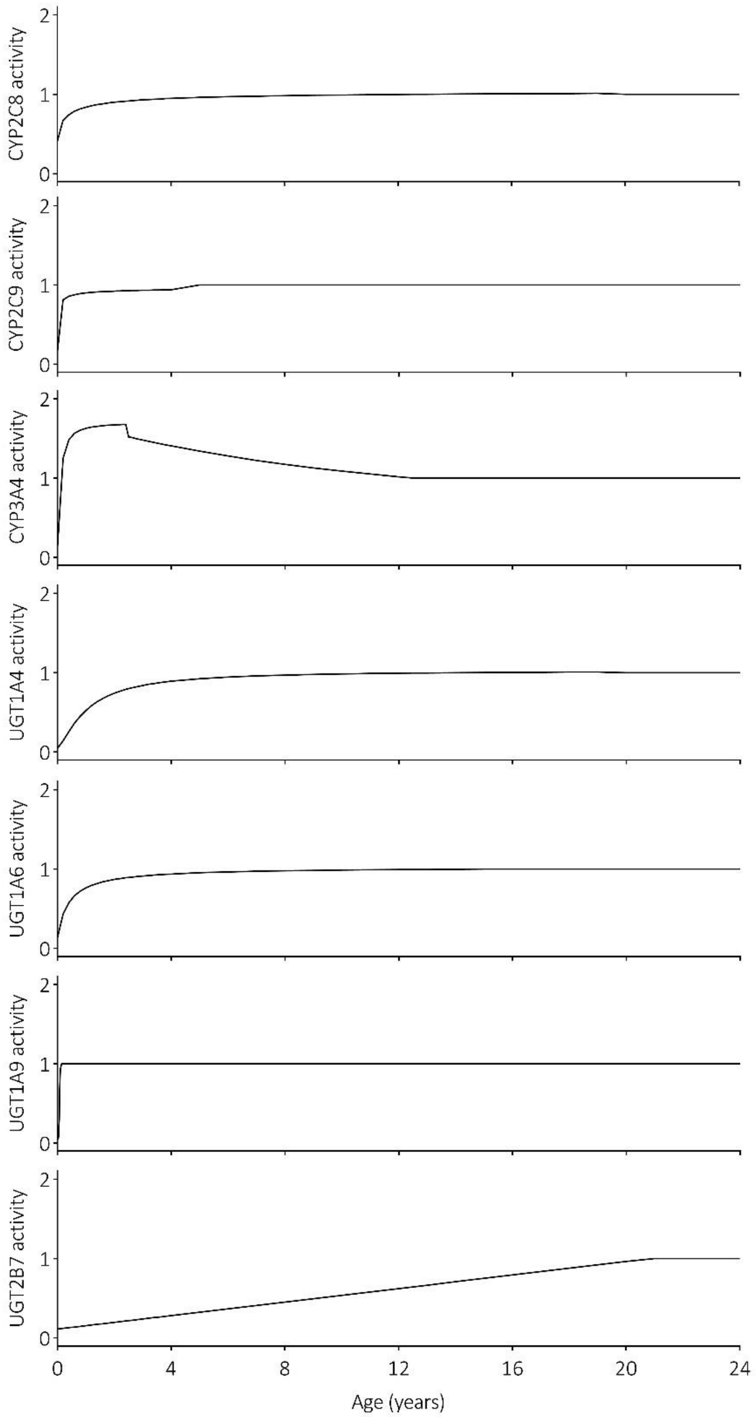

**Figure 1. CYP and UGT ontogeny profiles.** Equations: CYP2C8 activity below 20 years = 0.41 + (((1.053 - 0.41)*$Age^{0.68}$) / ($0.366^{0.68}$ + $Age^{0.68}$)); CYP2C9 activity below 5 years = 0.17 + (((0.98 - 0.17)*$Age^{0.53}$) / ($0.0157^{0.53}$ + $Age^{0.53}$)); CYP3A4 activity below 2.5 years = 0.15 + (((1.7 - 0.15)*$Age^{1.3}$) / ($0.1^{1.3}$ + $Age^{1.3}$)) and CYP3A4 activity above 2.5 years = 0.7 + 1*$exp^{-0.01*(Age-0.5)}$; UGT1A4 activity below 20 years = 0.05 + (((1.028 - 0.05)*$Age^{1.36}$) / ($1.042^{1.36}$ + $Age^{1.36}$)); UGT1A6 activity below 15 years = 0.142 + (((1.025 - 0.142)*$Age^{0.97}$) / ($0.411^{0.97}$ + $Age^{0.97}$)); UGT1A9 activity below 0.16 years = 0.0541 + (((1.012 - 0.0541)*$Age^{5.771}$) / ($0.07228^{5.771}$ + $Age^{5.771}$)); UGT2B7 activity below 21 years = 0.113 + 0.0425*Age.

#### 2. Search strategies

The PubMed database was searched for carbamazepine (CBZ) and valproic acid (VPA) adult and pediatric pharmacokinetic (PK) data. Both the drugname and brandname are included in the search queries. For CBZ the terms 'Carbamazepine' and 'Tegretol' were included and for VPA the terms 'Valproic Acid', 'Depakine', 'Depakote', 'Valproate', and 'VPA' Titles and abstracts of all search results were screened to check if actual PK data were provided in the publication. To extend our search strategy, the 'Similar articles' overview and references were checked to identify relevant articles which were missed with the initial search queries.

Search query for healthy volunteer PK studies:

((Pharmacokinet*[Title/Abstract]) AND (***DRUGNAME****[Title] OR ***BRANDNAME****[Title]) AND (Healthy[Title] OR volunteer*[Title] OR adult*[Title] OR subject*[Title] OR Man[Title] OR Men[Title] OR Woman[Title] OR Women[Title] OR male*[Title] OR female*[Title])) OR (("Pharmacokinetics of"[Title]) AND ("***DRUGNAME***"[Title] OR "***BRANDNAME***"[Title]) OR ("***DRUGNAME*** pharmacokinetics"[Title]))

CBZ → 176 results (01-08-2023)

VPA → 264 results (19-12-2023)

Search query for pediatric PK studies:

(Pharmacokinet*[Title/Abstract]) AND (***DRUGNAME****[Title] OR ***BRANDNAME****[Title]) AND (Infan*[Title] OR newborn*[Title] OR new-born*[Title] OR perinat*[Title] OR neonat*[Title] OR baby[Title] OR baby*[Title] OR babies[Title] OR prematur*[Title] OR preterm*[Title] OR toddler*[Title] OR minors[Title] OR minors*[Title] OR boy[Title] OR boys[Title] OR boyfriend[Title] OR boyhood[Title] OR girl*[Title] OR kid[Title] OR kids[Title] OR child[Title] OR child*[Title] OR children*[Title] OR schoolchild*[Title] OR schoolchild[Title] OR school child[Title] OR school child*[Title] OR adolescen*[Title] OR juvenil*[Title] OR youth*[Title] OR teen*[Title] OR under*age*[Title] OR pubescen*[Title] OR pediatrics[MeSH] OR pediatric*[Title] OR paediatric*[Title] OR peadiatric*[Title])

CBZ → 37 results (01-08-2023)

VPA → 82 results (19-12-2023)

#### 3. Pharmacokinetic studies

**Table 3.** Selected adult single-dose pharmacokinetic studies for carbamazepine model verification.

| | Study | N | Dose | Administration | Health status | Age range (years) | % females | Ref |
|---|---|---|---|---|---|---|---|---|
| Adult single dose | | | | | | | | |
| A | Barzaghi et al. 1987 | 7 | 400 mg | Oral | Healthy volunteers | 21-27 | 0 | (4) |
| B | Bedada et al. 2015 | 12 | 200 mg | Oral | Healthy volunteers | 25-30 | 0 | (5) |
| C | Bedada et al. 2016 | 12 | 200 mg | Oral | Healthy volunteers | 28-32 | 0 | (6) |
| D | Bedada et al. 2017 | 12 | 200 mg | Oral | Healthy volunteers | 25-33 | 0 | (7) |
| E | Bernus et al. 1994 | 6 | 600 mg | Oral | Healthy volunteers | 22-26 | 0 | (8) |
| F | Chan et al. 1985 | 6 | 400 mg | Oral | Healthy volunteers | 35.7 ± 8.3 | NS | (9) |
| G | Dalton et al. 1985 | 8 | 600 mg | Oral | Healthy volunteers | 24-35 | 0 | (10) |
| H | Gerardin et al. 1976 | 6 | 100, 200, and 600 mg | Oral | Healthy volunteers | NS | 0 | (11) |
| I | Maas et al. 1987 | 11 | 200 mg | Oral | Healthy volunteers | 21-30 | 54.5 | (12) |
| J | Marino et al. 2012 | 2 | 500 mg q12h, 100 mg | Oral, 10 min IV | Epileptic subjects | 41 ± 11 | 51.1 | (13) |
| K | Pynnönen et al. 1977 | 6 | 400 mg | Oral | Healthy volunteers | 21-22 | 0 | (14) |
| L | Pynnönen et al. 1978 | 8 | 400 mg (A) | Oral | Healthy volunteers | 23-30 | NS | (15) |
| M | Wada et al. 1978 | 9 | 200 mg | Oral | Healthy volunteers | NS | 0 | (16) |

Abbreviations: intravenously (IV), not specified (NS), number (N), reference (Ref), every 12 hours (q12h).

**Table 4.** Selected adult multi-dose pharmacokinetic studies for carbamazepine model verification.

| | Study | N | Dose | Administration | Health status | Age range (years) | % females | Ref |
|---|---|---|---|---|---|---|---|---|
| Adult multi dose | | | | | | | | |
| A | Burstein et al. 2000 | 8 | 100 mg bid d1-3, 200 mg bid d4-6, and 400 mg qd d7-21 | Oral | Healthy volunteers | 24-43 | 37.5 | (17) |
| B | Chan et al. 1985 | 10 | 200 mg bid d1-24 | Oral | Healthy volunteers | 40.2 ± 6.8 | NS | (9) |
| C | Gerardin et al. 1976 | 5 | 200 mg, after 96h 200 mg q24h d1-17 | Oral | Healthy volunteers | NS | 0 | (11) |
| D | Ji et al. 2013 | 12 | 200 mg qd d1-3, 200 mg bid d4-6, and 400 mg qd d7-21 | Oral | Healthy volunteers | 18-45 | 30.6 | (18) |
| E | Møller et al. 2000 | 12 | 100 mg bid d1-3, 200 mg bid d4-6, and 400 mg qd d7-21 | Oral | Healthy volunteers | 20-43 | 0 | (19) |
| F | Moreno et al. 2004 | 14 | 200 mg q8h, >1 month | Oral | Epileptic women | 21-44 | 100 | (20) |
| G | Patsalos et al. 1990 | 12 | 1200, 1400, 1600, and 1800 mg TID (8.00, 13.00 and 20.00) | Oral | Subjects with severe refractory epilepsy | 30-44 | 33.3 | (21) |
| H | Tolbert et al. 2015 | 23<br>41<br>10 | 178.24 mg q6h<br>163.48 mg q6h<br>168.28 mg q6h | 30 min IV<br>15 min IV<br>2-5 min IV | Types of epilepsy for which carbamazepine is indicated* | 20-65<br>23-68<br>23-53 | 47.8<br>53.7<br>60.0 | (22) |

*i.e., partial seizures with complex symptomatology [e.g., psychomotor, temporal lobe], generalized tonic–clonic seizures [i.e., grand mal], mixed seizure patterns that include one of the previous seizure types, or other partial or generalized seizures. Abbreviations: two times a day (bid), day (d), intravenously (IV), not specified (NS), number (N), every day (qd), reference (Ref), every x hours (qxh).

**Table 5.** Selected pediatric pharmacokinetic single-dose studies for carbamazepine model verification

| | Study | N | Dose | Administration | Health status | Age range | % females | Ref |
|---|---|---|---|---|---|---|---|---|
| Pediatric single dose | | | | | | | | |
| A | Bano et al. 1986 | 6 | 9.3 ± 0.87 mg/kg | Oral tablets | Healthy children | 7-14 y | 33.3 | (23) |
| B | Bertilsson et al. 1980 | 2<br>1 | 2 mg/kg<br>3 mg/kg | Gelatin capsule<br>Gelatin capsule | Children with psychomotor epilepsy | 10-11 y<br>13 y | 50<br>100 | (24) |
| C | Mackintosh et al. 1987 | 6 | 5 mg/kg | Oral suspension | New-born infants with convulsions | GA 32-42w<br>PNA 2-16d | 50 | (25) |
| D | Rey et al. 1979 | 7<br>5 | 17 mg/kg<br>19 mg/kg | Oral suspension<br>Oral suspension | Newborns hospitalized for epilepsy<br>Children hospitalized for epilepsy | 20.9 d<br>5.1 y | NS<br>NS | (26) |
| E | Singh et al. 1996 | 10 | 10 mg/kg | Oral suspension | Term neonates in the NICU with 2 or more seizures | 0-7 d | NS | (27) |

Abbreviations: days (d), gestational age (GA), number (N), neonatal intensive care unit (NICU), postnatal age (PNA), reference (Ref), weeks (w), year (y).

**Table 6.** Selected pediatric pharmacokinetic multi-dose studies for carbamazepine model verification

| | Study | N | Dose | Administration | Health status | Age range (years) | % females | Ref |
|---|---|---|---|---|---|---|---|---|
| Pediatric multi dose | | | | | | | | |
| A | Bertilsson et al. 1980 | 3 | Custom | Oral tablets | Children with psychomotor epilepsy | 10-13 | 66.6 | (24) |
| B | Camfield et al. 1992 | 15<br>1<br>1 | 6.75 mg/kg q12h (2.2-10.3 mg/kg)[a]<br>200 mg q8h<br>200 mg at 9 AM, and 300 mg at 2 and 8 PM | Oral tablets | Children with epilepsy | 11±3 (5-16) | 47.7 | (28) |
| C | Carlsson et al. 2005 | 52 | 7.35 mg/kg q12h (3.2-16.05 mg/kg)[a] | Oral suspension, IR and CR tablets | Pediatric patients with epilepsy | 2-21 (2-18) | 36.8 | (29) |
| D | Cornaggia et al. 1993 | 15 | 7.56 mg/kg at 9 AM, 3 and 9 PM (4.3-13.7 mg/kg)[a] | Oral tablets | Pediatric patients with EPS/CPS, secondarily generalized seizures, and GTCS | 6-14 | 53.3 | (30) |
| E | Eeg-Olofsson et al. 1990 | 21 | 9.55 mg/kg q12h (10.0-35.3 mg/kg/day)[a] | Oral tablets | Pediatric patients with partial or GTCS | 4-13 | 47.6 | (31) |
| F | Furlanut et al. 1984 | 45 | 6.3 mg/kg q8h | Oral tablets | Pediatric patients with GTCS, EPS and CPS | 0.417-17 | 44.4 | (32) |
| G | Hartley et al. 1991 | 21 | 7.60 mg/kg q12h | Oral tablets | Pediatric patients with epilepsy | 6.97-15.67 | 33.3 | (33) |
| H | Hartley et al. 1991a | 12 | 10 mg/kg q12h | Oral tablets | Pediatric patients with GTCS or CPS | 6.5-15 | 25.0 | (34) |
| I | Reith et al. 2000 | 6 | 13.85 mg/day (6.925 mg q12h) | Oral tablets | Children on carbamazepine treatment | 4-10 | 42.9 | (35) |
| | | 11 | | | | 10-13 | | |
| | | 4 | | | | 13-19 | | |
| J | Thakker et al. 1992 | 12 | 233.3 mg 5h, 183.3 mg 4.8h, 183.3mg 3.3h, and 233.3 mg 10.9h | Oral tablets | Pediatric patients with chronic seizure disorder | 8-17 | 0.25 | (36) |

[a]Average dose (range) Abbreviations: Controlled-release (CR), Generalised tonic-clonic seizures (GTCS), Elementary partial epileptic seizures (EPS), Complex partial epileptic seizures (CPS), Immediate-release (IR), every x hour (qxh), number (N), reference (Ref).

**Table 7.** Selected adult pharmacokinetic studies for valproic acid immediate-release model verification

| | Study | N | Dose (pure VPA) | Administration | Health status | Age range (years) | % females | Ref |
|---|---|---|---|---|---|---|---|---|
| 1.1 Adult single dose | | | | | | | | |
| A | Bialer et al. 1985 | 6 | 1000 mg **(867.7)** | Oral (Sodium VPA) | Healthy volunteers | 20-35 | 0 | (37) |
| G | Cheng et al. 2007 | 1 | 6.66 mg/kg **(5.78)** | Oral (Sodium VPA) | AML patient | NS | NS | (38) |
| B | Chun et al. 1980 | 9 | 250 mg | Oral Capsule | Healthy volunteers | 21-28 | 0 | (39) |
| C | Georgoff et al. 2018 | 44 | 140 mg/kg **(121.5)**<br>130 mg/kg **(112.8)**<br>15 mg/kg **(13.0)** | 1 hour IV infusion | Healthy volunteers | 18-60 | 11.4 | (40) |
| D | Gugler et al. 1977 | 6 | 600 mg **(520.6)** | Oral – Enteric coated | Healthy volunteers | 23-26 | 16.7 | (41) |
| E | Nitsche and Mascher 1982 | 6 | 1000 mg **(867.7)** | 5 min IV bolus | Healthy volunteers | 19-31 | 0 | (42) |
| F | Perucca et al. 1978 | 6 | 800 mg **(694.2)**<br>800 mg **(694.2)** | Oral (Sodium VPA)<br>5 min IV bolus | Healthy volunteers | 22-38 | 0 | (43) |
| 1.2 Adult multi dose | | | | | | | | |
| A | Addison et al. 2000 | 7 | 200 mg q12h **(173.5)** | Oral (Sodium VPA) | Healthy volunteers | 19-30 | 0 | (44) |
| B | Cheng et al. 2007 | 1 | 6.66 mg/kg q8h **(5.78)** | Oral (Sodium VPA) | AML patient | NS | NS | (38) |
| C | Hussein et al. 1994 | 18 | 750 mg **(650.81)** followed by 250 mg q6h **(216.94)** | 1 hour IV infusion | Healthy volunteers | 18-34 | 0 | (45) |

Abbreviations: acute myeloid leukemia (AML), intravenous (IV), not specified (NS), number (N), reference (Ref), every x hours (qxh).

**Table 8.** Selected pediatric pharmacokinetic studies for valproic acid immediate release model verification

| | Study | N | Dose (pure VPA) | Administration | Health status | Age range (years) | % females | Ref |
|---|---|---|---|---|---|---|---|---|
| **1.1 Pediatric single dose** | | | | | | | | |
| A | Alfonso et al. 2000 | 1<br>1 | 10 mg/kg<br>25 mg/kg | 30 min IV infusion<br>30 min IV infusion | Hypomelanosis<br>Epileptic | 21 days<br>28 days | 100<br>0 | (46) |
| B | Cloyd et al. 1983 | 12 | 11.2 ± 6.9 mg/kg **(9.72)** | PO sodium VPA syrup | Children taking antiepileptics | 1-14 | 59 | (47) |
| C | Hall et al. 1983 | 1<br>25 | 10.4 mg/kg **(9.02)**<br>11.0 ± 3.7 mg/kg **(9.54)** | PO sodium VPA[a] (syrup or capsule) | Pediatric patients with seizures | 11<br>0.25-21 | NS<br>32 | (48) |
| D | Visudtibhan et al. 2011 | 11 | 18.4 mg/kg **(15.97)** | 6.13 min IV sodium VPA | Thai pediatric patients with varying underlying diseases | 1-15 | 72.7 | (49) |
| E | Williams et al. 2012 | 10 | 15.0 mg/kg **(13.02)** | 5 or 10 min IV sodium VPA | Epileptic | 1-17 | NS | (50) |
| F | Brachet-Lierman & Demarquez 1977 | 1 | 100 mg/kg **(86.8)** | PO dipropylacetate | Epileptic | 3 days | NS | (51) |
| **1.2 Pediatric multi dose** | | | | | | | | |
| A | Chiba et al. 1985 | 21 | 13.9 ± 3.55 mg/kg q12h **(12.06)** | PO sodium VPA (12 uncoated tablet/ 9 syrup) | Japanese epileptic children | 3.6-15.2 | 52.4 | (52) |
| B | Cloyd et al. 1983 | 25 | 22.95 ± 13.1 mg/kg q12h **(19.91)** | PO sodium VPA syrup | Children taking antiepileptics | 1-14 | 59 | (47) |
| C | Gal et al. 1988 | 5 | 20-25 mg/kg loading dose **(19.52)**,<br>5-10 mg/kg q12h maintenance **(6.51)** | PO Sodium VPA syrup | Neonates with intractable seizures | 6-31 days | NS | (53) |
| D | Hall et al. 1983 | 1<br>25 | 12.6 mg/kg q12h **(10.93)**<br>16.4 ± 8.2 mg/kg q12h **(14.23)** | PO sodium VPA[a] (syrup or capsule) | Pediatric patients with seizures | 11<br>0.25 – 21 | NS<br>32 | (48) |
| E | Herngren et al. 1991 | 7 | 14.83 ± 3.94 mg/kg q12h **(12.87)**<br>16.5 mg/kg q8h **(14.32)** | PO sodium VPA (solution/ mixture) | Generalized epilepsies | 0.5 – 1.5 | 29 | (54) |
| F | Lundberg et al. 1982 | 7<br>4 | 15.14 ± 3.53 mg/kg q12h **(13.14)**<br>21.75 ± 10.80 mg/kg q8h **(18.87)** | PO sodium VPA tablets | Epileptic | 6-13<br>4.5-9.5 | 28.6<br>50 | (55) |
| G | Su et al. 2011 | 4 | 12.5 ± 3.5 mg/kg q12h **(10.84)** | PO syrup/capsules | Refractory solid/CNS tumors | 3-21 | 61 | (56) |
| H | Zhao et al. 2016 | 75<br>95 | 19.63 ± 7.29 mg/kg q24h **(17.03)**<br>23.67 ± 9.21 mg/kg q24h **(20.54)** | PO sodium VPA | Chinese epilepsy patients | 0-3<br>3-25 | 44<br>45.3 | (57) |

[a]It is assumed that sodium VPA is administered. Abbreviations: central nervous system (CNS), intravenous (IV), not specified (NS), number (N), reference (Ref), every x hours (qxh).

**Table 9.** Selected adult pharmacokinetic studies for valproic acid extended-release formulation model verification

| | Study | N | Dose (pure VPA) | Administration | Health status | Age range (years) | % females | Ref |
|---|---|---|---|---|---|---|---|---|
| 1.1 Adult single dose | | | | | | | | |
| A | Dulac et al. 2005 | 22 | 500 mg | PO SR tablets | Healthy volunteers | 18-34 | 0 | (58) |
| B | Dutta et al. 2004A | 15 | 500 mg | PO divalproex sodium ER tablets | Healthy volunteers | 19-53 | 12.8 | (59) |
| C | Dutta et al. 2004B | 16 | 500 mg | PO divalproex sodium ER tablets | Healthy volunteers | 18-50 | 37.5 | (60) |
| D | Qiu et al. 2003 | 15 | 500 mg | PO divalproex sodium CR tablets | Healthy volunteers | NS | NS | (61) |
| 1.2 Adult multi dose | | | | | | | | |
| A | Cawello et al. 2012 | 16 | Day 1-3: 150 mg q12h **(130.16)**<br>Day 4-17: 300 mg q12h **(260.31)** | PO SR tablets (Ergenyl Chrono 300) | Healthy volunteers | 22-43 | 0 | (62) |
| B | Dulac et al. 2005 | 22 | 500 mg q12h | PO SR tablets | Healthy volunteers | 18-34 | 0 | (58) |
| C | Dutta et al. 2002 | 35 | 1000 mg q24h<br>1500 mg q24h | PO divalproex sodium ER tablets | Healthy volunteers | 19-55 | 36.1 | (63) |
| D | Qiu et al. 2003 | 15 | 1000 mg q24h | PO divalproex sodium CR tablets | Healthy volunteers | NS | NS | (61) |

Abbreviations: controlled-release (CR), extended-release (ER), intravenously (IV), not specified (NS), number (N), reference (Ref), sustained-release (SR), every x hours (qxh).

**Table 10.** Selected pediatric pharmacokinetic studies for valproic acid extended-release model verification

| | Study | N | Dose (pure VPA) | Administration | Health status | Age (years) | % females | Ref |
|---|---|---|---|---|---|---|---|---|
| 1.2 Pediatric multi dose | | | | | | | | |
| A | Dutta et al. 2004D | 14<br>12 | 19.9 ± 8.54 mg/kg q24h<br>14.4 ± 5.64 mg/kg q24h | PO divalproex sodium ER tablets | Pediatric patients with migraine, bipolar disorder, ADHD or seizures | 8-11<br>13-17 | NS | (64) |
| B | Herranz et al. 2006 | 48 | 11.3 ± 3.15 mg/kg q12h **(9.81)**<br>22.55 ± 6.25 mg/kg q24h **(19.57)** | PO SR valproate tablets (depakine chrono) | Pediatric patients with epilepsy | 4.83-13.83 | 60.4 | (65) |
| | | 11 | 11.95 ± 2.4 mg/kg q12h **(10.37)** | | | 5.25-13.83 | 45.5 | |
| | | 11 | 29.1 ± 8.0 mg/kg q24h **(25.25)** | | | 4.83-13.83 | 54.5 | |
| | | 11 | 23.7 ± 5.7 mg/kg q24h **(20.56)** | | | 4.83-13.42 | 81.8 | |

Abbreviations: attention-deficit hyperactivity disorder (ADHD), intravenously (IV), not specified (NS), number (N), reference (Ref), every x hours (qxh).

### 4. Model verification

#### 4.1 Model verification workflow and model performance assessment

Before model application, model performance need to be assessed via model verification. The verification workflow has been published earlier following a pragmatic approach (66, 67). In short, the CBZ and VPA PBPK models, as well as their metabolites, CBZE and 4-ene-VPA, respectively, were verified in the adult population. Simulations upon intravenous (IV) and oral (PO) administration (single and multidose) were compared with published PK data (Table S3, 4, 7 and 9). All simulations were conducted with 10 virtual subjects in 10 trials (i.e., n=100), unless stated otherwise below. In case steady state CBZ concentrations were obtained while no specific sampling day was indicated in the study, it was decided to run the simulations for 31 days and to compare the observed data with the predicted data from the last day.

Of note, for simulations where a 'Custom dosing scheme' or IV administration was applicable (also in simulations with children), CBZ was included in Simcyp as an 'Inhibitor' with CBZE as the 'Inhibitor Metabolite'. As CBZE elimination is partly parameterized as an Oral In Vivo Clearance (Table S1) was it incompatible with the custom dosing functionality when incorporated as 'Sub 1 Met'. The default Alfentanil model was applied as substrate, because there is no interaction between alfentanil and CBZ. Hence, alfentanil has no impact on predicted CBZ concentrations.

Next, simulations with pediatrics receiving IV or PO administrations were conducted and evaluated with clinical data (Table S5, 6, 8 and 10). CYP3A4 is the main metabolizing enzyme of CBZ, whereas VPA is mainly metabolized by UGT's. For CYP2C8, CYP2C9, CYP3A4, UGT1A4, UGT1A6, UGT1A9 and UGT2B7 are ontogeny profiles included (Figure S1), while for CYP3A5, UGT1A8 and UGT1A10 no ontogeny is included (i.e., fraction of adult activity is 1 for all ages). The 'redefine subjects over time' function was activated.

Specific CBZ virtual study designs which were not straight forward:

- For Patsalos et al. (21), four separate simulations were conducted where either 1200, 1400, 1600 or 1800 mg per day was administered orally. Then, plasma concentration-time data were combined to obtain a mean profile of the 4 different dosages to reflect the observed data (n=400).
- For Bertilsson et al. (24), for each separate child, an oral custom dosing scheme was simulated which were as follows: Subject A – Day 1: 2 mg/kg once, day 5-8: 2.62 mg/kg every 12 hours (q12h), day 26-146: 2.43 mg/kg q12h. Subject B – Day 1: 3 mg/kg once, day 5-8: 4.13 mg/kg q12h, day 9-12: 2.08 mg/kg at 9 AM and 4.13 mg/kg at 9 PM, day 13-153: 2.08 mg/kg q12h. Subject C – Day 1: 3 mg/kg once, day 5-38: 1.84 mg/kg at 9 AM and 3.71 mg/kg at 9 PM, day 39-162: 3.45 mg/kg q12h. Individual PK parameters are obtained from these simulations.

- For Camfield et al. (28), three separate simulations are conducted. One to derive PK parameters from (i.e., 6.75 mg/kg q12h) and two to derive individual plasma concentration-time profiles from to match to pediatric subjects (i.e., 200 mg q8h which is 'subject 1' and 200 mg at 9 AM, and 300 mg at 2 and 8 PM which is 'subject 2').

Specific VPA virtual study designs which were not straight forward:

- For Herranz et al. 2006 (65), observed data from VPA administration once daily in the morning and evening are represented in one simulation. As well as the 1 month and 4 months VPA concentration measurements are combined to one timepoint for each study.

Predictive performance of PBPK models was evaluated by 1) calculating the ratio of predicted-to-observed (p/o) PK parameters, and by 2) a visual predictive check (VPC). Ratios within 0.5 to 2-fold range were considered acceptable. For a visual check, observed plasma concentration-time profiles were extracted from literature, digitalized with WebPlotDigitizer v4.6, and compared to predicted plasma concentration-time curves.

4.2 Model verification results

For CBZ and CBZE predictions, the adult p/o PK ratios are shown in Figure S2, the pediatric ratios are shown in Figure S3, whereas for VPA the ratios upon IR administration are provided in Figure S4 and ratios upon ER administration in Figure S5. The percentages of p/o CBZ and CBZE PK ratios that fell within the 2-, 1.5-, and 1.25-fold range are provided in Table S11 and for VPA in Table S12. The VPCs for CBZ and CBZE predictions in adults are shown in Figure S6-7, in pediatrics in Figure S8-9, whereas for VPA IR in Figure S10-12 and for VPA ER in Figure S13-14.

**Table 11. Carbamazepine (CBZ) and carbamazepine-10,11-epoxide (CBZE) ratios within 2-, 1.5-, and 1.25-fold range.**

| | CBZ | | | | CBZE | | | |
|---|---|---|---|---|---|---|---|---|
| | N | 2-fold | 1.5-fold | 1.25-fold | N | 2-fold | 1.5-fold | 1.25-fold |
| Adult single | 53 | 66% | 40% | 23% | 17 | 76% | 35% | 24% |
| Adult multi | 40 | 100% | 85% | 53% | 28 | 86% | 54% | 39% |
| Pediatric single | 28 | 82% | 43% | 25% | 0 | | | |
| Pediatric multi | 39 | 100% | 90% | 62% | 14 | 79% | 79% | 71% |

Table 12. Valproic acid (VPA) ratios within 2-, 1.5-, and 1.25-fold range.

| | VPA IR | | | | VPA ER | | | |
|---|---|---|---|---|---|---|---|---|
| | N | 2-fold | 1.5-fold | 1.25-fold | N | 2-fold | 1.5-fold | 1.25-fold |
| Adult single | 33 | 93.9% | 90.9% | 63.6% | 18 | 88.9% | 61.1% | 44.4% |
| Adult multi | 8 | 100% | 100% | 87.5% | 16 | 87.5% | 87.5% | 68.8% |
| Pediatric single | 18 | 66.7% | 61.1% | 61.1% | 0 | | | |
| Pediatric multi | 33 | 87.9% | 48.5% | 39.4% | 10 | 100% | 60.0% | 50.0% |

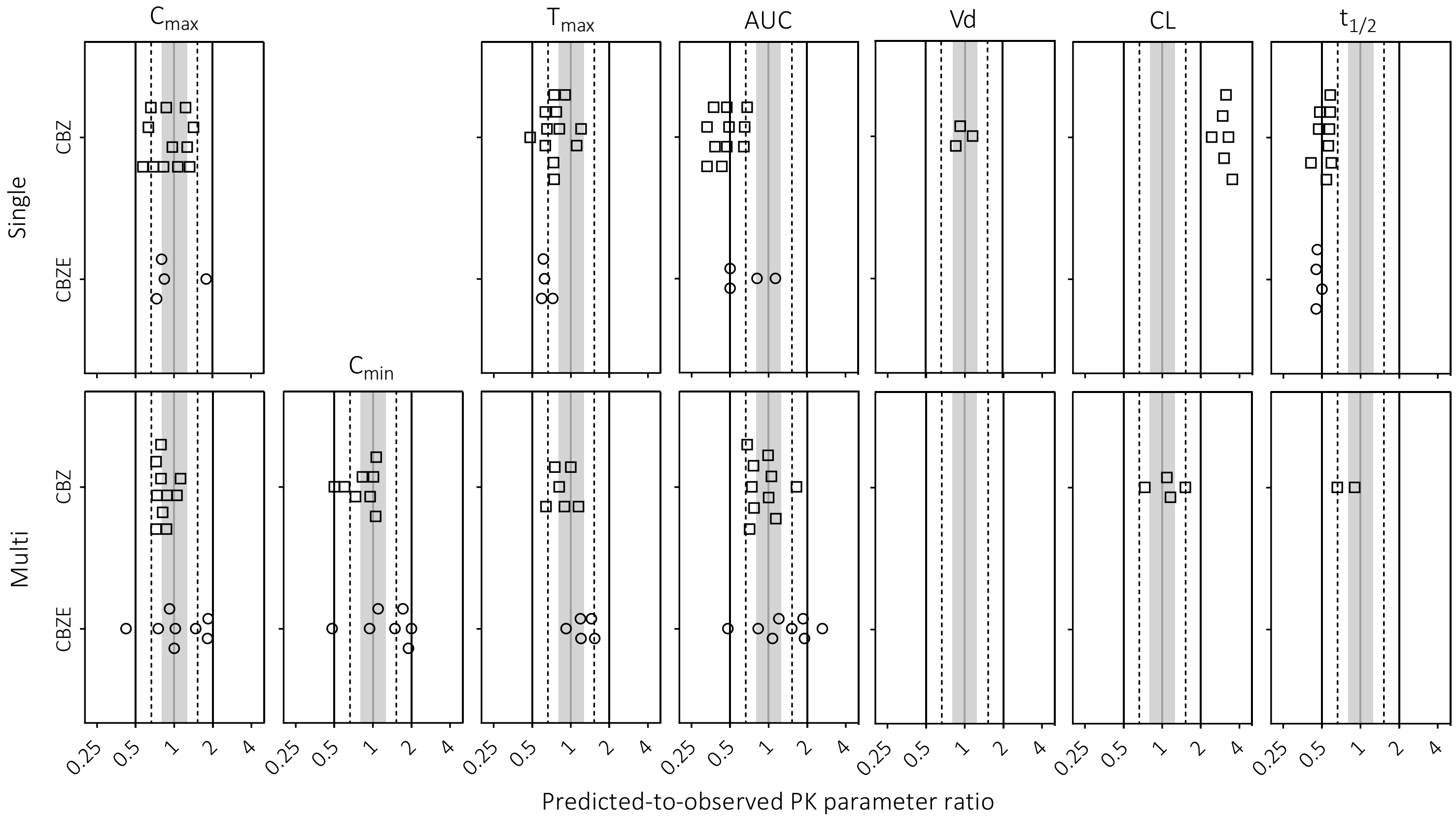

**Figure 2. Predicted-to-observed ratios of the maximum concentration ($C_{max}$), minimum concentration ($C_{min}$), time to peak concentration ($T_{max}$), area under the curve (AUC), volume of distribution (Vd), clearance (CL), and half-life ($t_{1/2}$) for carbamazepine single- and multi-dose studies in adults.** Single symbols represent a predicted-to-observed ratio of a single pharmacokinetic study. The black lines represent the 2-fold range, the dashed lines the 1.5-fold range, the gray shaded area represent the 1.25-fold range and the gray line represents the unity line.

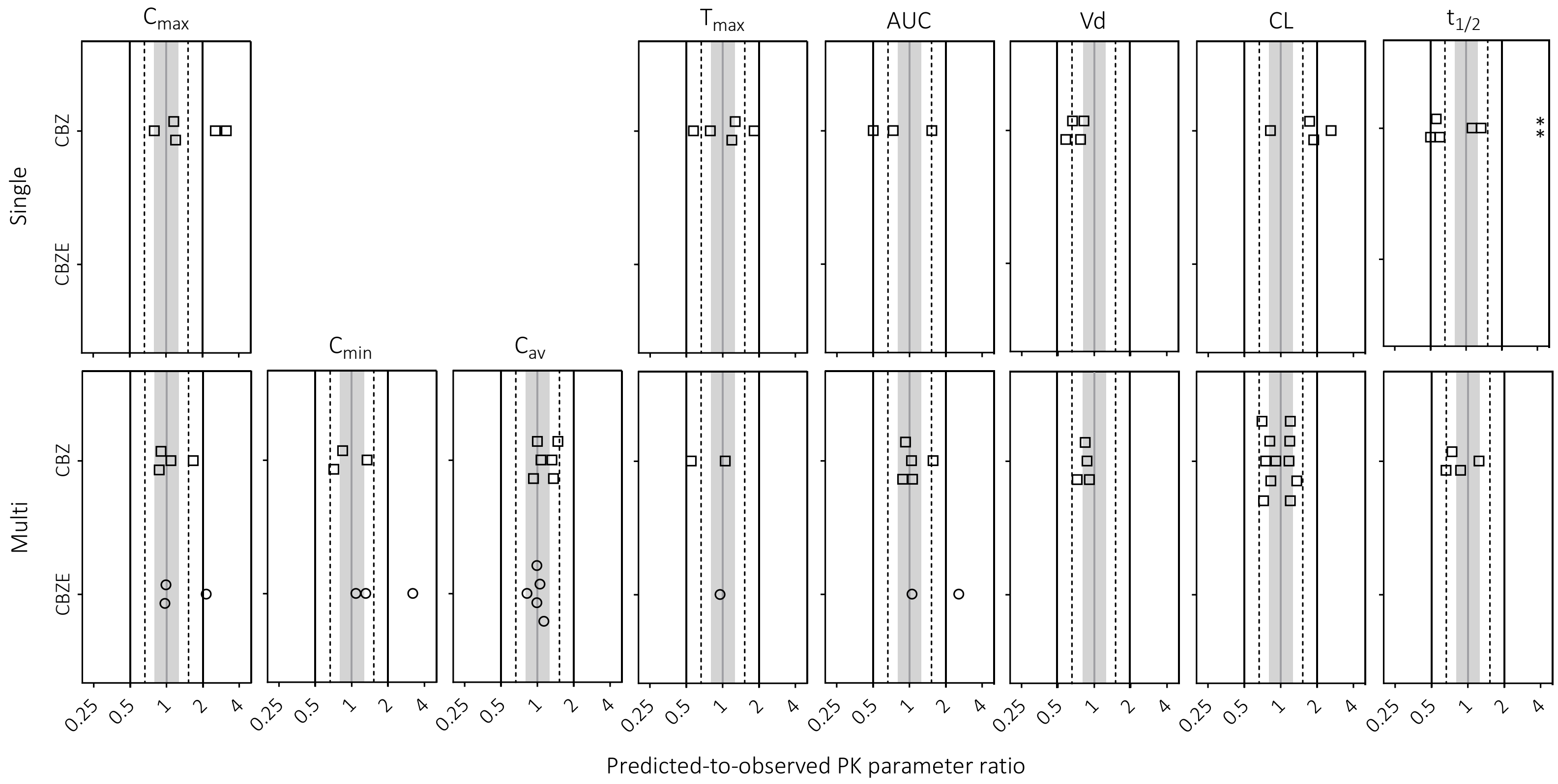

**Figure 3. Predicted-to-observed ratios of the maximum concentration ($C_{max}$), minimum concentration ($C_{min}$), average concentration ($C_{av}$), time to peak concentration ($T_{max}$), area under the curve (AUC), volume of distribution (Vd), clearance (CL), and half-life ($t_{1/2}$) for carbamazepine single- and multi-dose studies in pediatrics.** Single symbols represent a predicted-to-observed ratio of a single pharmacokinetic study. The black lines represent the 2-fold range, the dashed lines the 1.5-fold range, the gray shaded area represent the 1.25-fold range and the gray line represents the unity line.

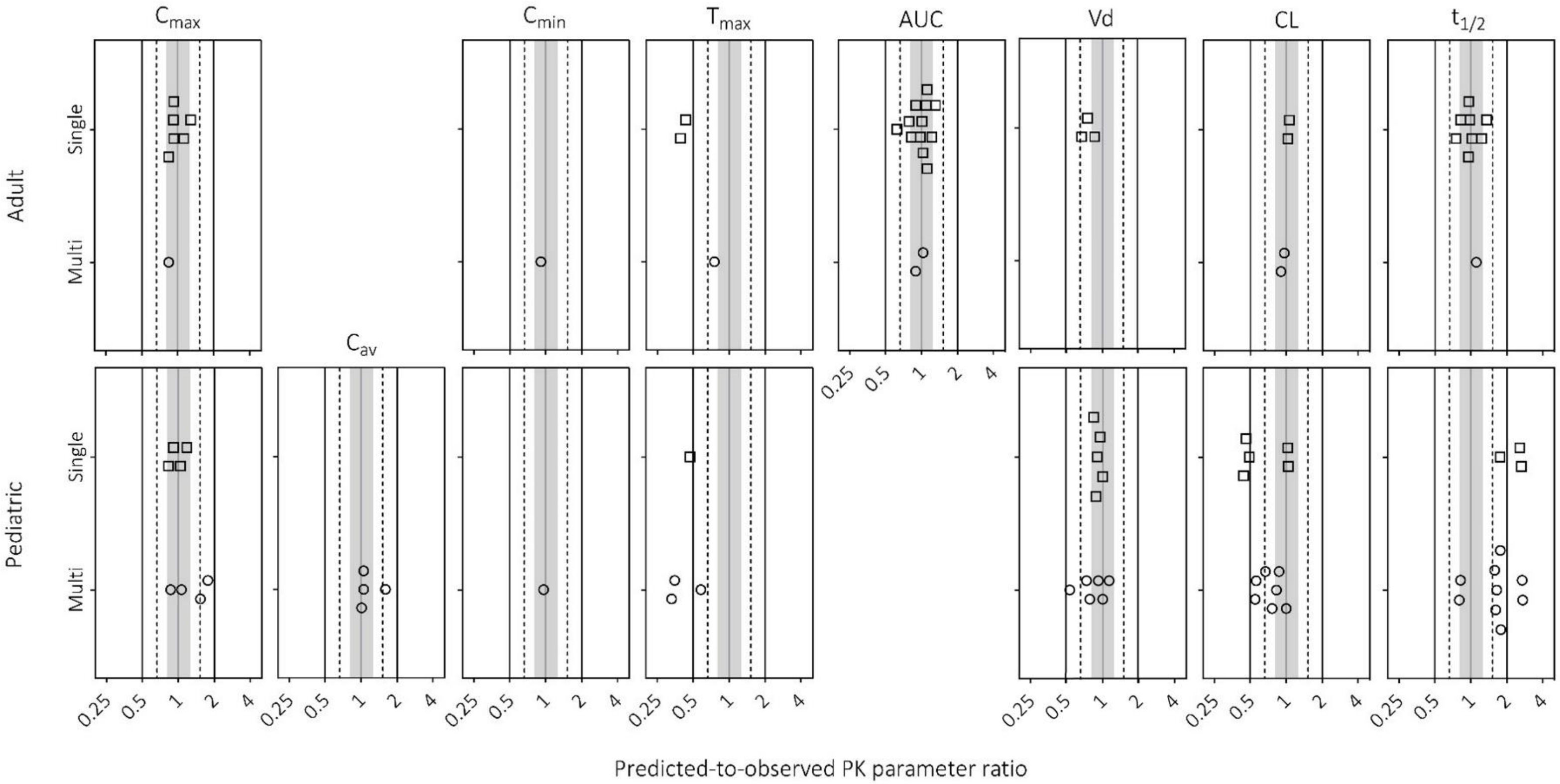

**Figure 4. Predicted-to-observed ratios of the maximum concentration ($C_{max}$), average concentration ($C_{av}$), minimum concentration ($C_{min}$), time to peak concentration ($T_{max}$), area under the curve (AUC), volume of distribution (Vd), clearance (CL), and half-life ($t_{1/2}$) for valproic acid immediate release formulation single- and multi-dose studies in adults and pediatrics.** Single symbols represent a predicted-to-observed ratio of a single pharmacokinetic study. The black lines represent the 2-fold range, the dashed lines the 1.5-fold range, the gray shaded area represent the 1.25-fold range and the gray line represents the unity line.

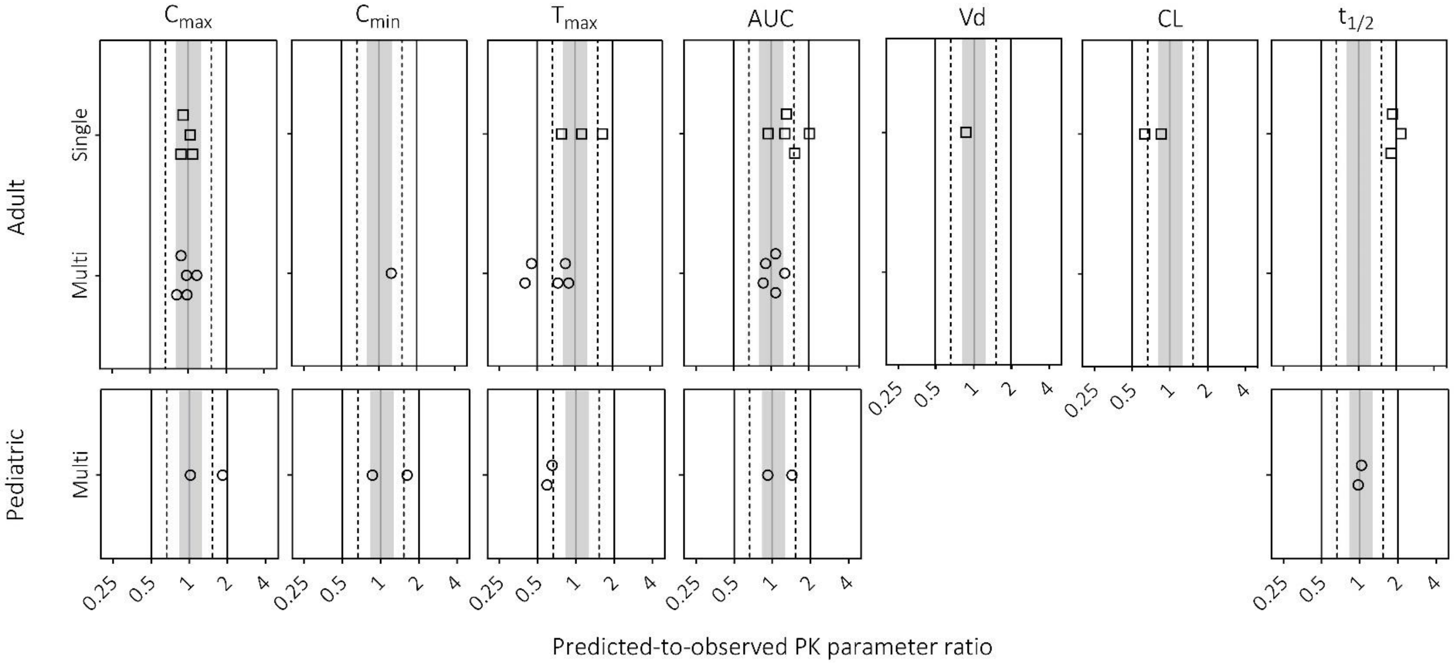

**Figure 5. Predicted-to-observed ratios of the maximum concentration ($C_{max}$), minimum concentration ($C_{min}$), time to peak concentration ($T_{max}$), area under the curve (AUC), volume of distribution (Vd), clearance (CL), and half-life ($t_{1/2}$) for valproic acid extended-release formulations single- and multi-dose studies in adults and pediatrics.** Single symbols represent a predicted-to-observed ratio of a single pharmacokinetic study. The black lines represent the 2-fold range, the dashed lines the 1.5-fold range, the gray shaded area represent the 1.25-fold range and the gray line represents the unity line.

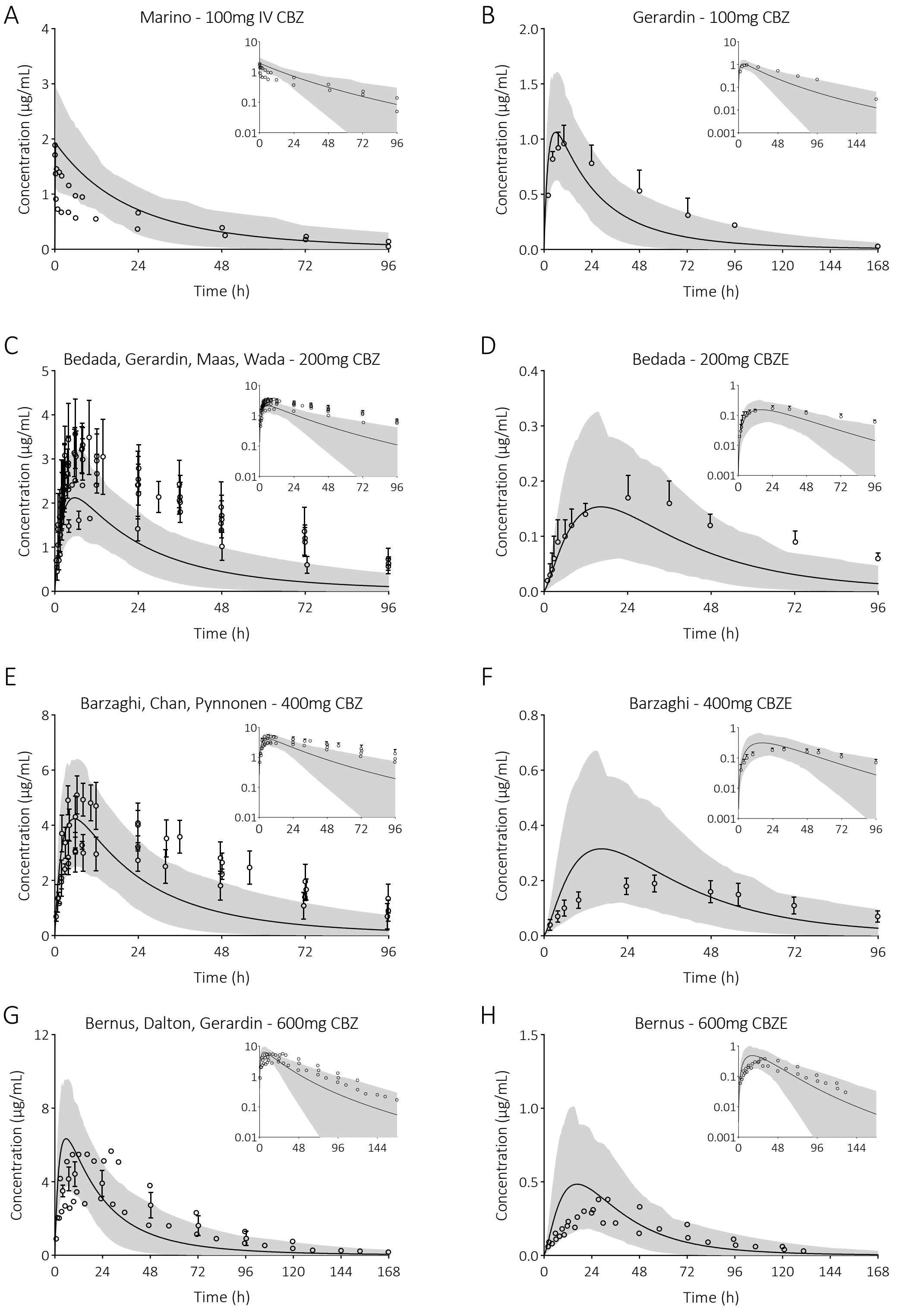

Figure 6. **Prediction of carbamazepine (CBZ) and carbamazepine-10,11-epoxide (CBZE) plasma concentration-time profiles in adults upon single dose administration.** The solid line is the predicted mean of the simulated population and the shaded area represents the 5$^{th}$ to 95$^{th}$ percentile of the virtual population. Open circles are the observed data: A (13), B (11), C (5-7, 11, 12, 16), D (5), E (4, 9, 14, 15), F (4), G (8, 10, 11), and H (8).

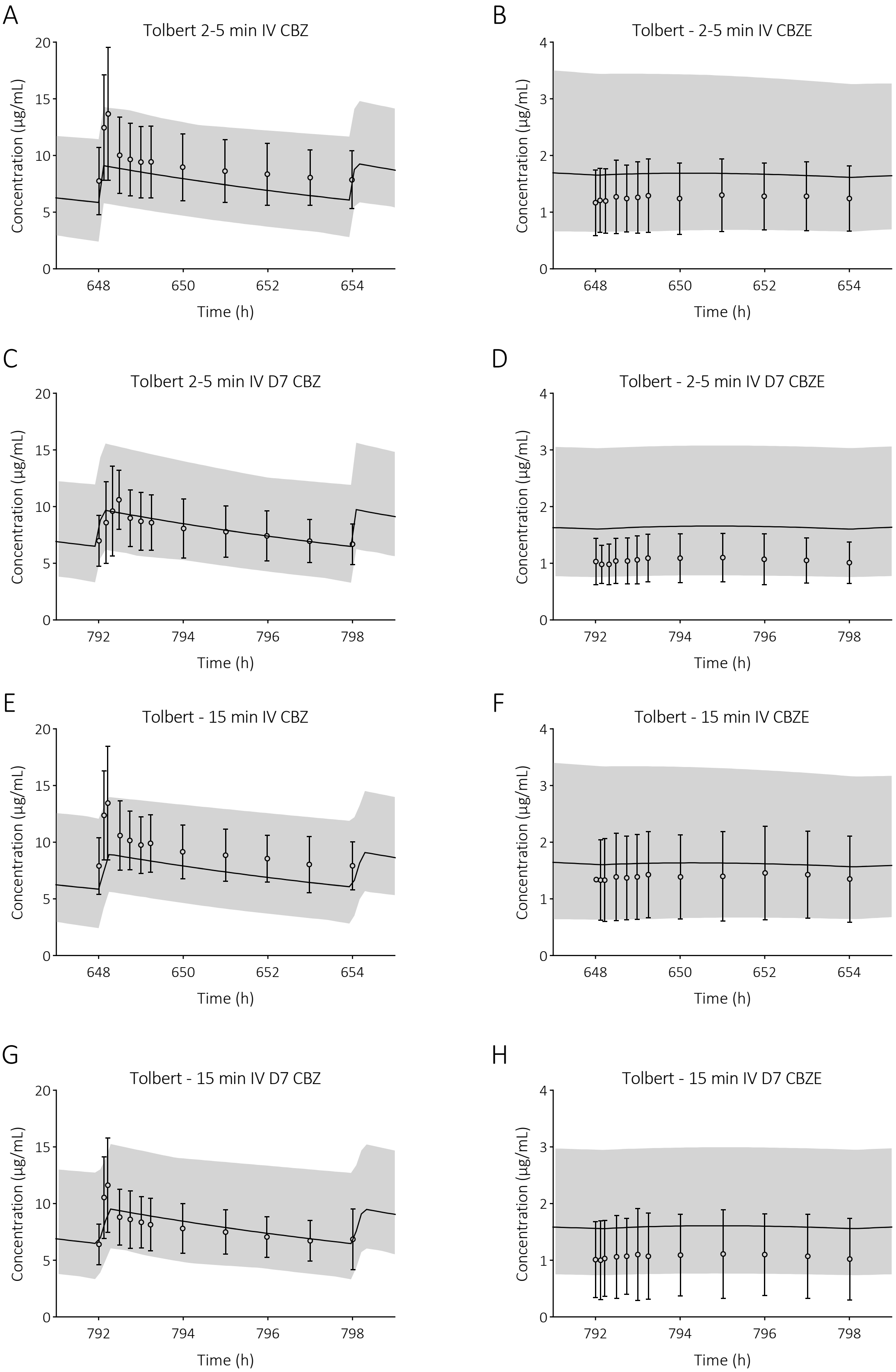

A
Tolbert 2-5 min IV CBZ
B
Tolbert - 2-5 min IV CBZE
C
Tolbert 2-5 min IV D7 CBZ
D
Tolbert - 2-5 min IV D7 CBZE
E
Tolbert - 15 min IV CBZ
F
Tolbert - 15 min IV CBZE
G
Tolbert - 15 min IV D7 CBZ
H
Tolbert - 15 min IV D7 CBZE
Concentration (µg/mL)
Time (h)
0
5
10
15
20
1
2
3
4
648
650
652
654
792
794
796
798

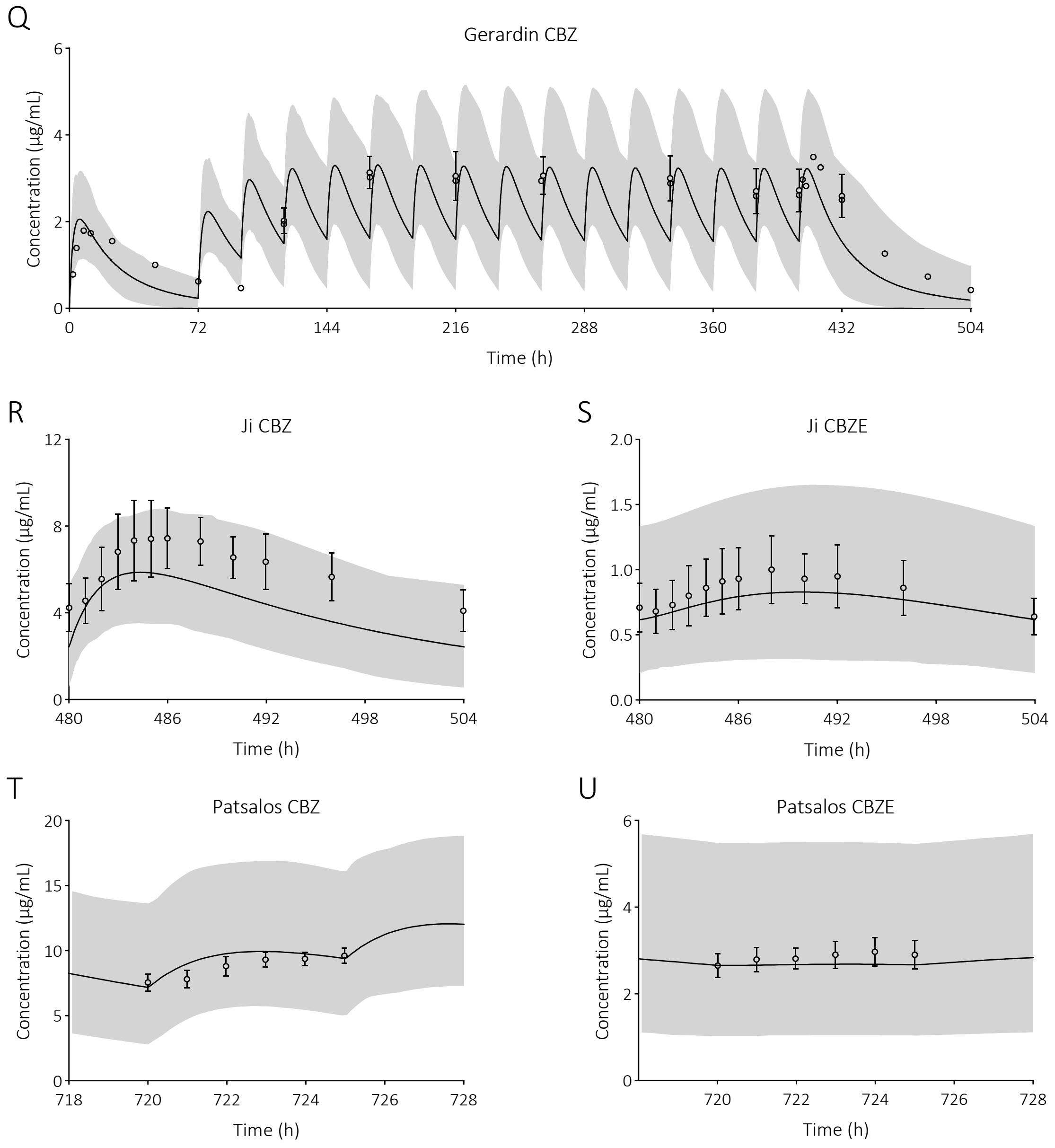

**Figure 7. Prediction of carbamazepine (CBZ) and carbamazepine-10,11-epoxide (CBZE) plasma concentration-time profiles in adults upon multi dose administration.** The solid line is the predicted mean of the simulated population and the shaded area represents the 5th to 95th percentile of the virtual population. Open circles are the observed data: A-L (22), M & N (17, 19), O & P (9), Q (11), R & S (18), and T & U (21).

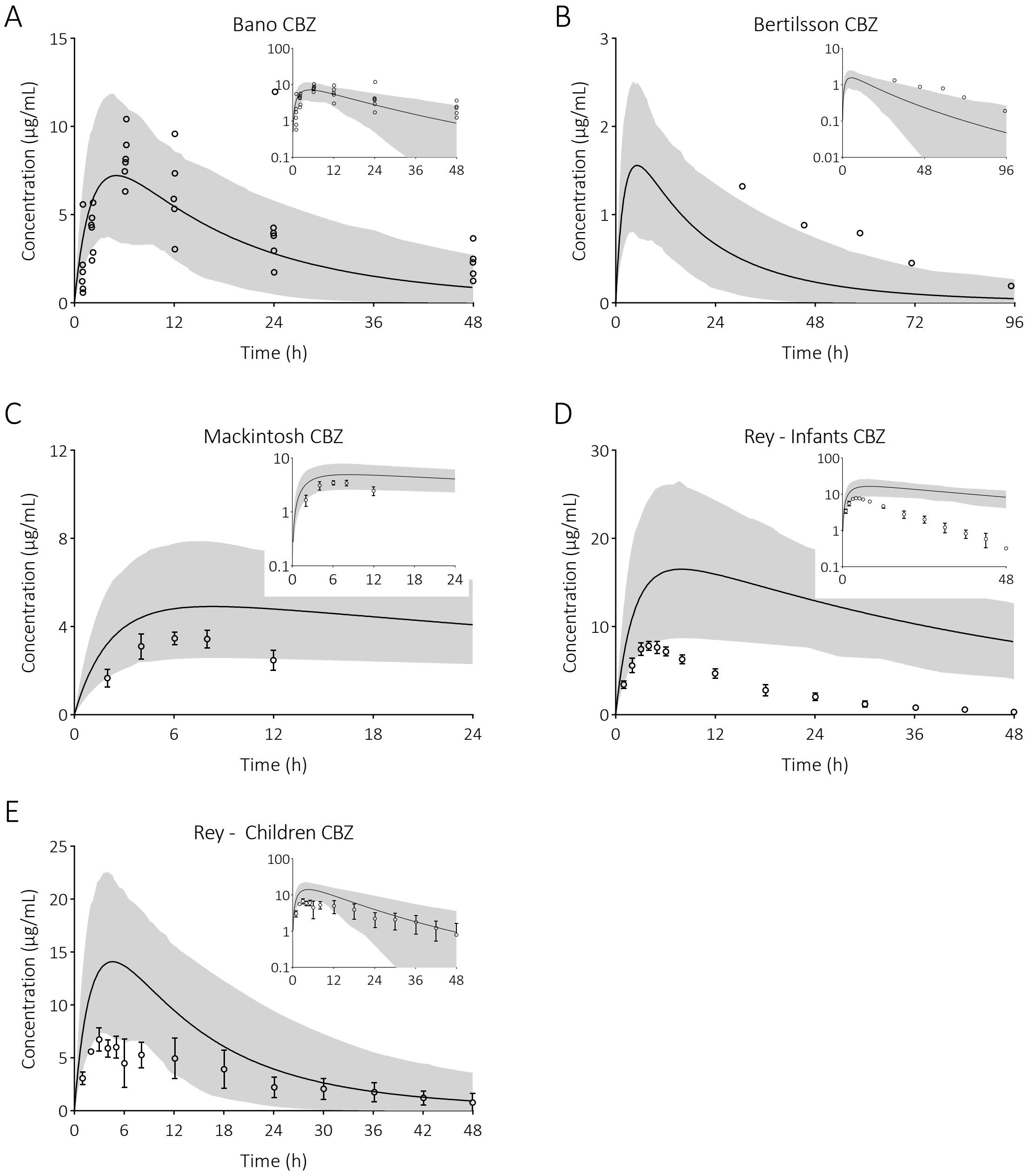

**Figure 8. Prediction of carbamazepine (CBZ) and plasma concentration-time profiles in paediatric individuals upon single dose administration.** The solid line is the predicted mean of the simulated population and the shaded area represents the 5$^{th}$ to 95$^{th}$ percentile of the virtual population. Open circles are the observed data: A (23), B (24), C (25), and D & E (26).

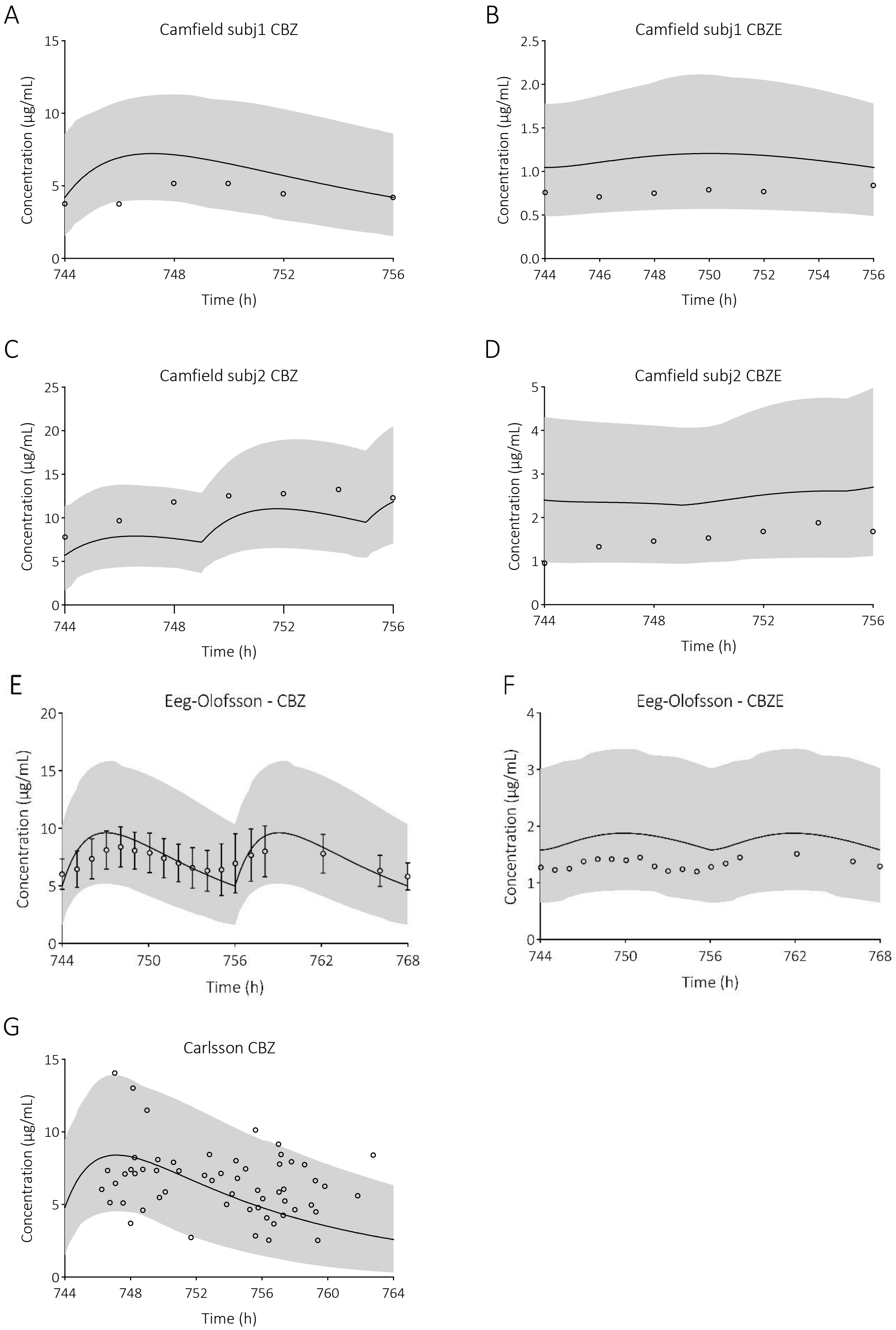
A
Camfield subj1 CBZ
Concentration (µg/mL)
0
5
10
15
744
748
752
756
Time (h)
B
Camfield subj1 CBZE
Concentration (µg/mL)
0.0
0.5
1.0
1.5
2.0
2.5
744
746
748
750
752
754
756
Time (h)
C
Camfield subj2 CBZ
Concentration (µg/mL)
0
5
10
15
20
25
744
748
752
756
Time (h)
D
Camfield subj2 CBZE
Concentration (µg/mL)
0
1
2
3
4
5
744
748
752
756
Time (h)
E
Eeg-Olofsson - CBZ
Concentration (µg/mL)
0
5
10
15
20
744
750
756
762
768
Time (h)
F
Eeg-Olofsson - CBZE
Concentration (µg/mL)
0
1
2
3
4
744
750
756
762
768
Time (h)
G
Carlsson CBZ
Concentration (µg/mL)
0
5
10
15
744
748
752
756
760
764
Time (h)

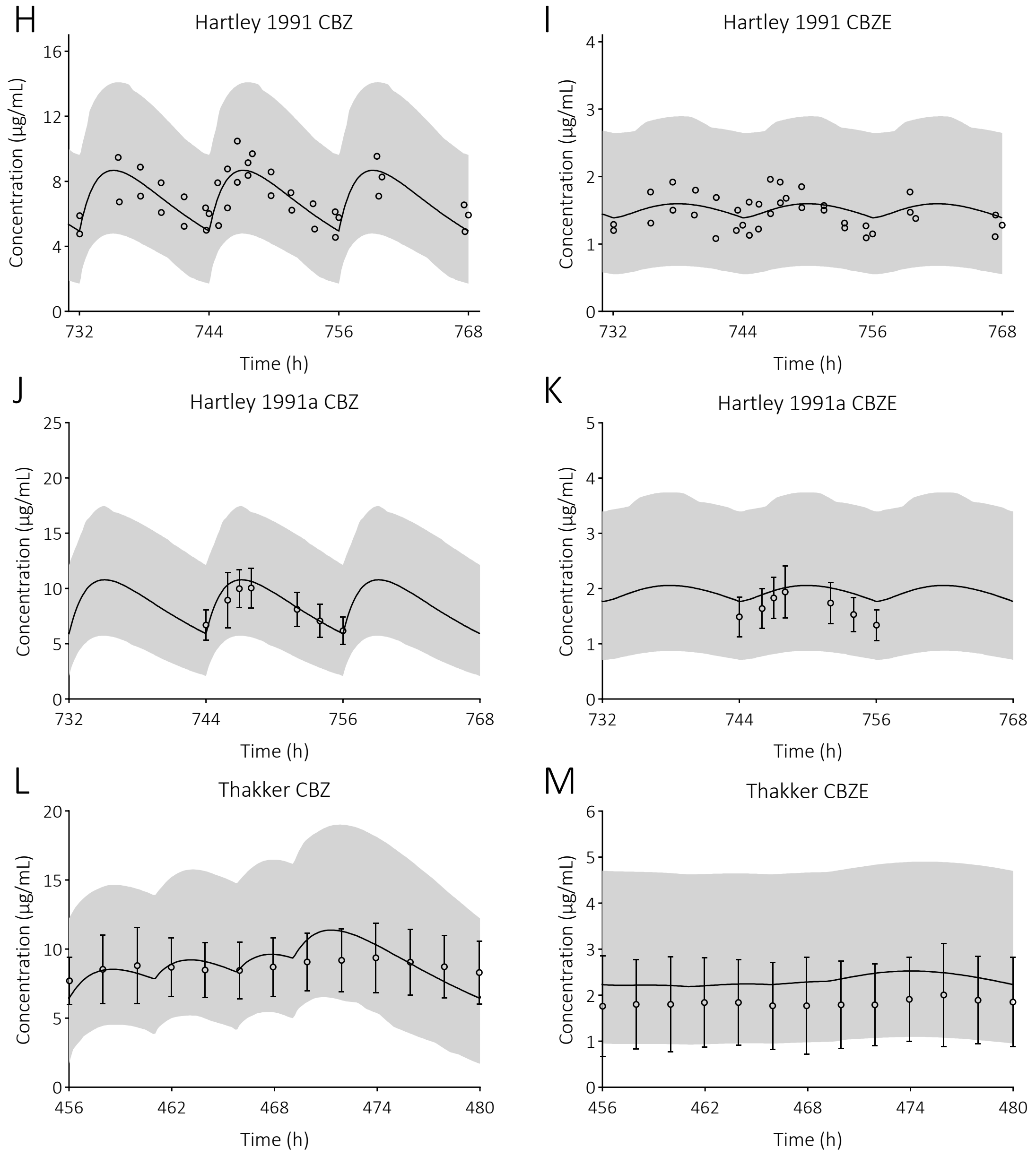

**Figure 9. Prediction of carbamazepine (CBZ) and carbamazepine-10,11-epoxide (CBZE) plasma concentration-time profiles in paediatric individuals upon multi dose administration.** The solid line is the predicted mean of the simulated population and the shaded area represents the 5$^{th}$ to 95$^{th}$ percentile of the virtual population. Open circles are the observed data: A-D (28), E & F (31), G (29), H & I (33), J & K (34), and L & M (36).

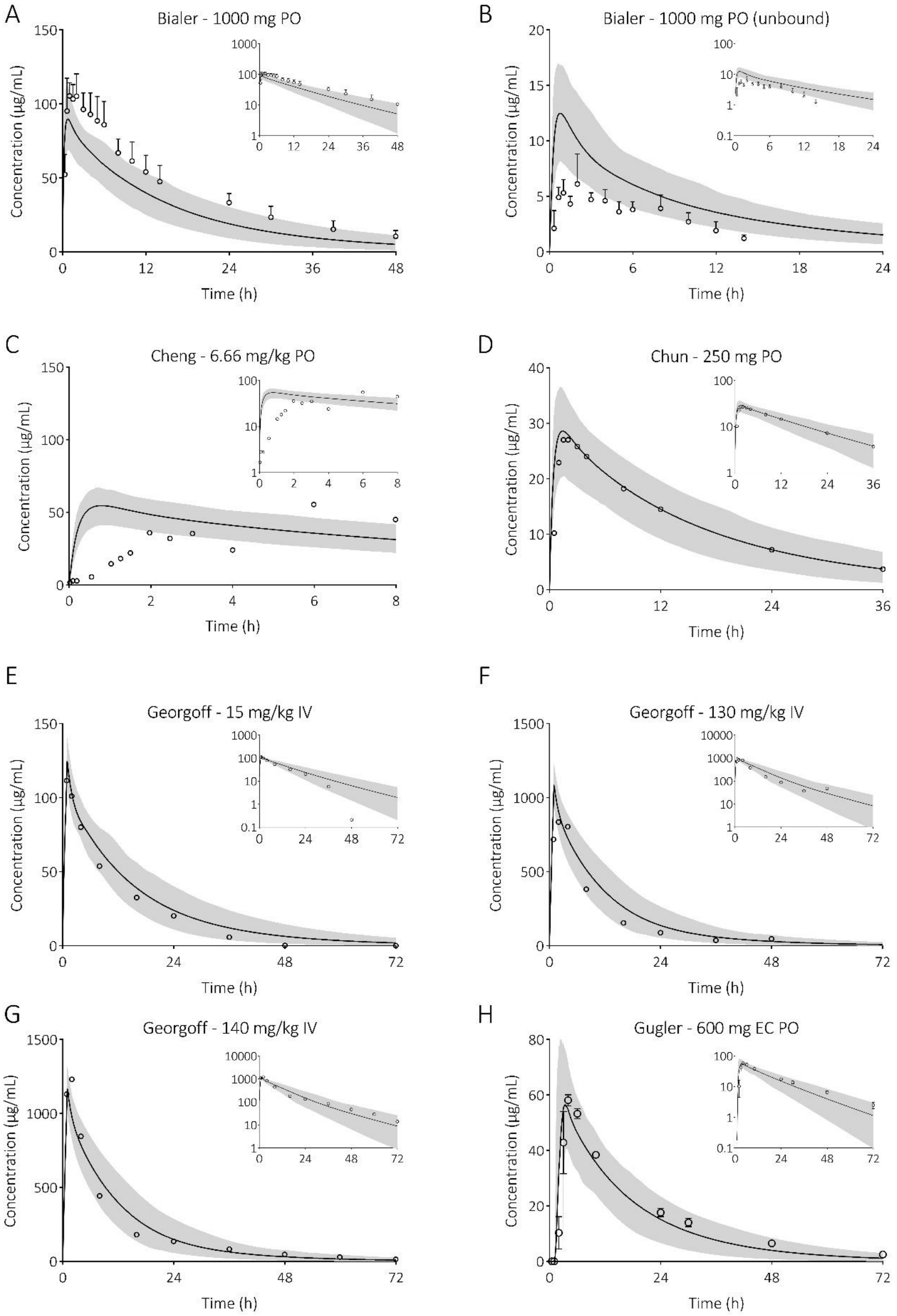
A
Bialer - 1000 mg PO
Concentration (µg/mL)
Time (h)
B
Bialer - 1000 mg PO (unbound)
Concentration (µg/mL)
Time (h)
C
Cheng - 6.66 mg/kg PO
Concentration (µg/mL)
Time (h)
D
Chun - 250 mg PO
Concentration (µg/mL)
Time (h)
E
Georgoff - 15 mg/kg IV
Concentration (µg/mL)
Time (h)
F
Georgoff - 130 mg/kg IV
Concentration (µg/mL)
Time (h)
G
Georgoff - 140 mg/kg IV
Concentration (µg/mL)
Time (h)
H
Gugler - 600 mg EC PO
Concentration (µg/mL)
Time (h)

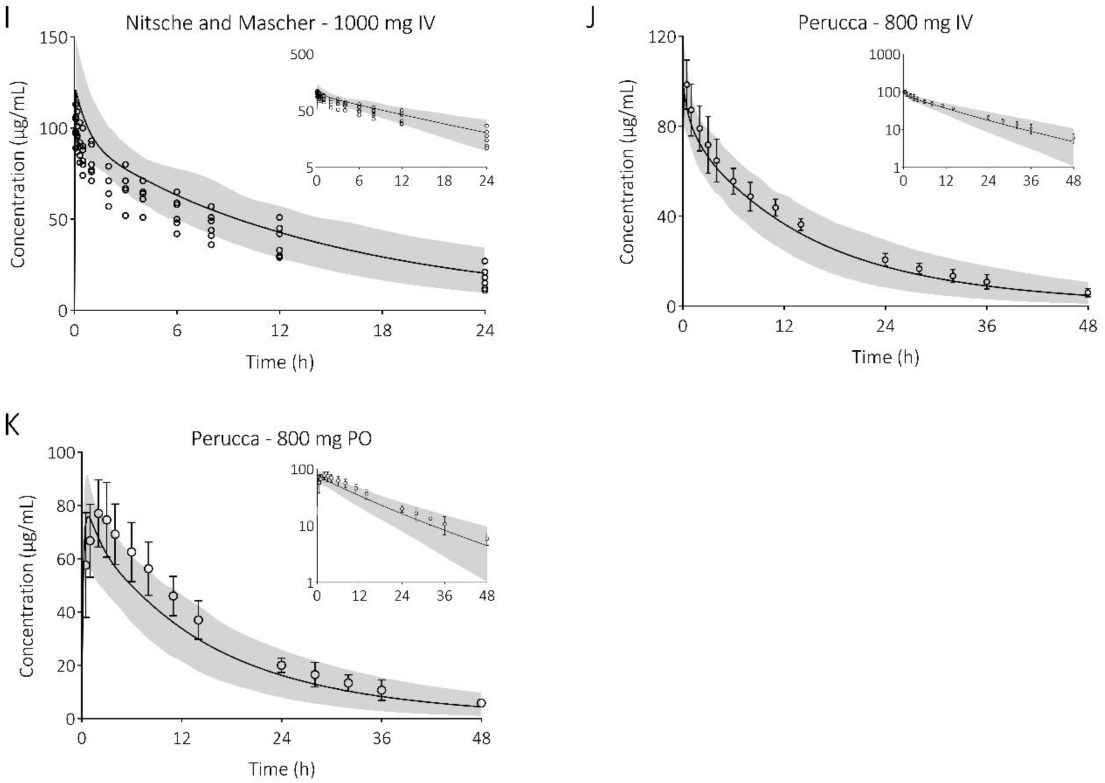

**Figure 10. Prediction of (unbound) valproic acid plasma concentration-time profiles in adults upon single-dose immediate-release administration.** The solid line is the predicted mean of the simulated population and the shaded area represents the 5$^{th}$ to 95$^{th}$ percentile of the virtual population. Open circles are the observed data: A & B (37), C (38), D (39), E-G (40), H (41), I (42), and J & K (43).

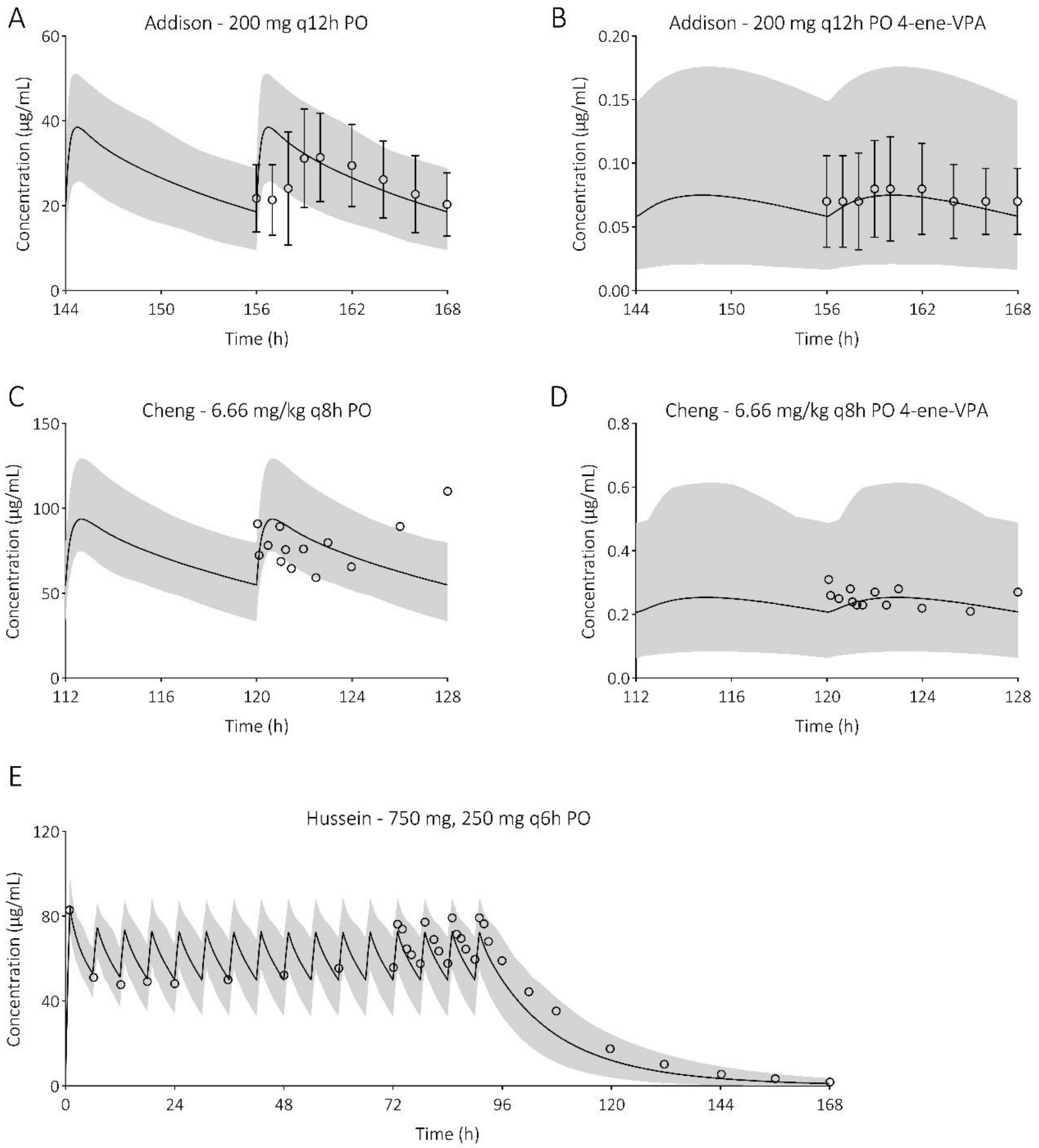

**Figure 11. Prediction of valproic acid (VPA) and 4-ene-VPA plasma concentration-time profiles in adults upon multi-dose immediate-release administration.** The solid line is the predicted mean of the simulated population and the shaded area represents the 5$^{th}$ to 95$^{th}$ percentile of the virtual population. Open circles are the observed data: A & B (44), C & D (38), and E (45).

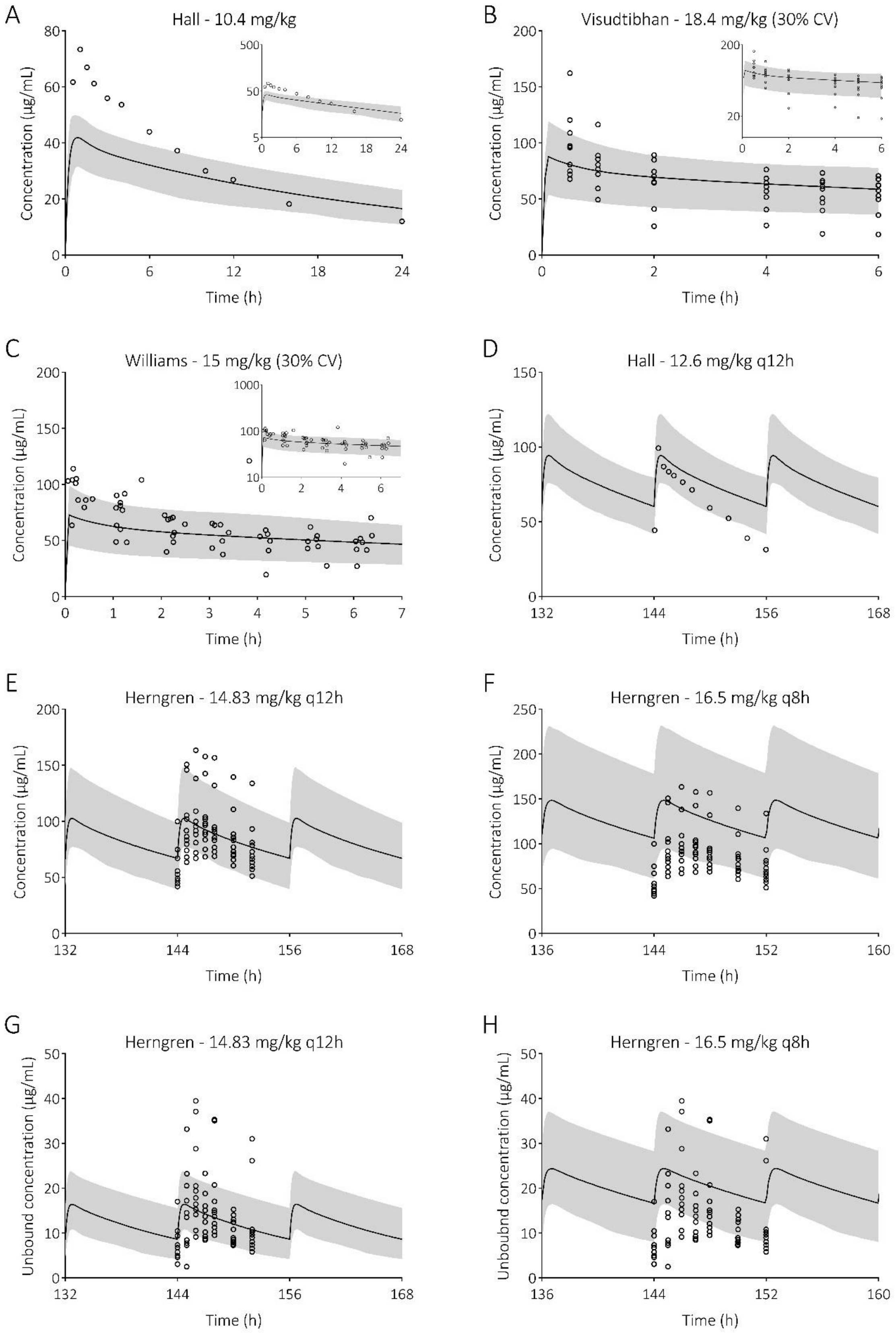
A
Hall - 10.4 mg/kg
Concentration (µg/mL)
Time (h)
B
Visudtibhan - 18.4 mg/kg (30% CV)
Concentration (µg/mL)
Time (h)
C
Williams - 15 mg/kg (30% CV)
Concentration (µg/mL)
Time (h)
D
Hall - 12.6 mg/kg q12h
Concentration (µg/mL)
Time (h)
E
Herngren - 14.83 mg/kg q12h
Concentration (µg/mL)
Time (h)
F
Herngren - 16.5 mg/kg q8h
Concentration (µg/mL)
Time (h)
G
Herngren - 14.83 mg/kg q12h
Unbound concentration (µg/mL)
Time (h)
H
Herngren - 16.5 mg/kg q8h
Unboubnd concentration (µg/mL)
Time (h)

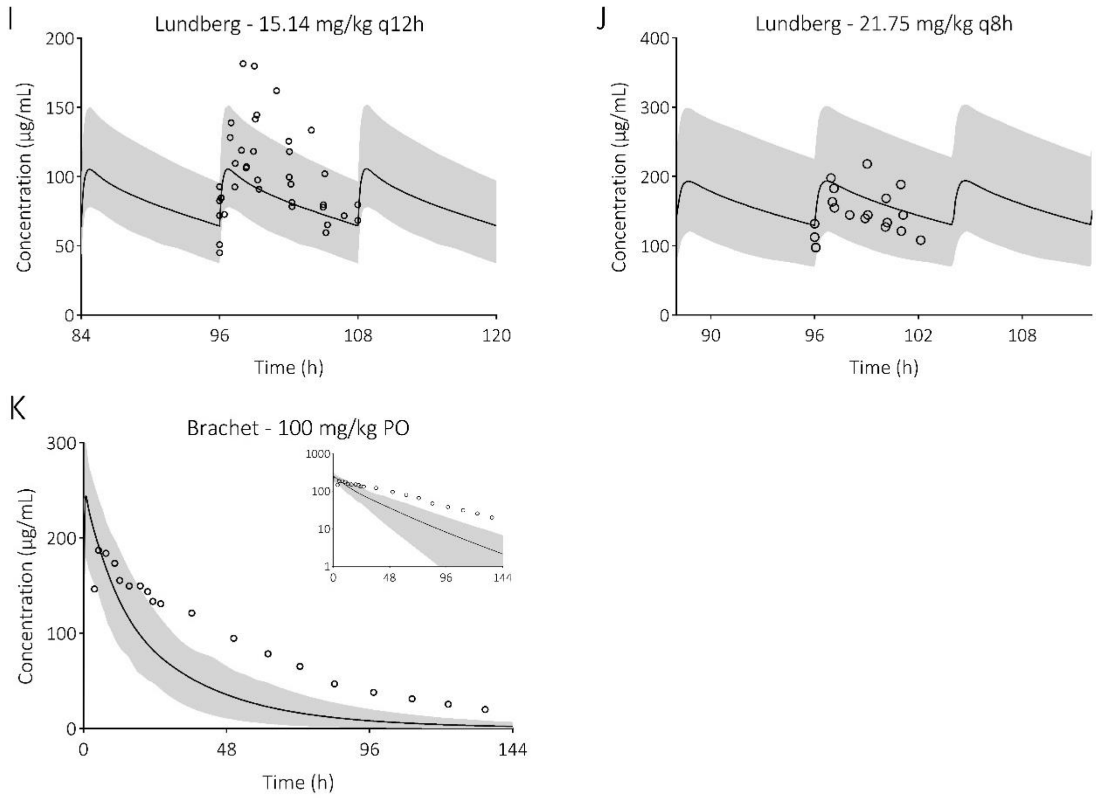

**Figure 12. Prediction of valproic acid (VPA) plasma concentration-time profiles in pediatrics upon single- and multi-dose immediate release administration.** The solid line is the predicted mean of the simulated population and the shaded area represents the 5th to 95th percentile of the virtual population. Open circles are the observed data: A (48), B (49), C (50), D (48), E-H (54), I & J (55), and K (51).

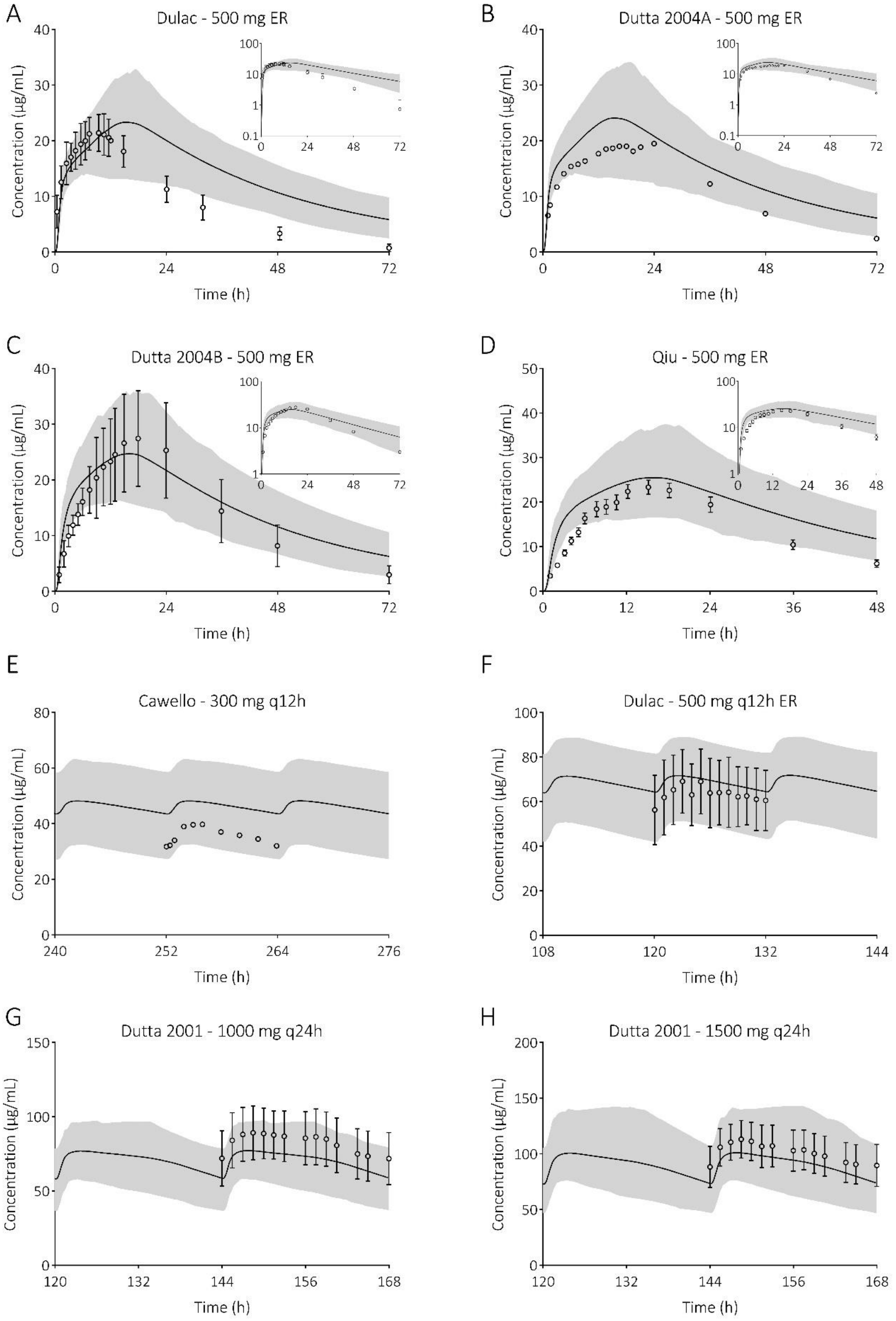

**Figure 13. Prediction of valproic acid (VPA) plasma concentration-time profiles in adults upon single- and multi-dose extended-release administration.** The solid line is the predicted mean of the simulated population and the shaded area represents the 5th to 95th percentile of the virtual population. Open circles are the observed data: A (58), B (59), C (60), D (61), E (62), F (58), and G & H (63).

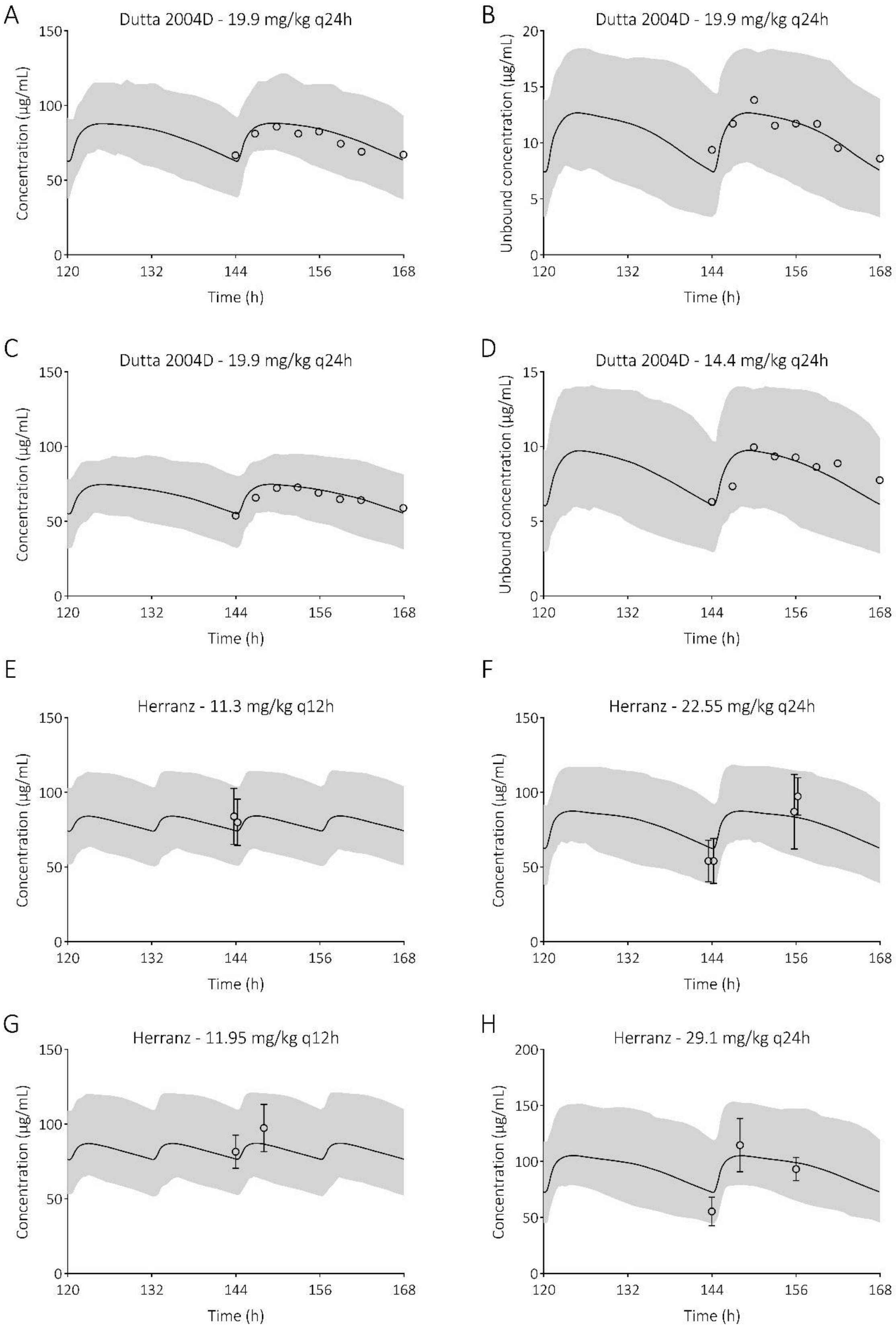
A
Dutta 2004D - 19.9 mg/kg q24h
Concentration (µg/mL)
Time (h)
B
Dutta 2004D - 19.9 mg/kg q24h
Unbound concentration (µg/mL)
Time (h)
C
Dutta 2004D - 19.9 mg/kg q24h
Concentration (µg/mL)
Time (h)
D
Dutta 2004D - 14.4 mg/kg q24h
Unbound concentration (µg/mL)
Time (h)
E
Herranz - 11.3 mg/kg q12h
Concentration (µg/mL)
Time (h)
F
Herranz - 22.55 mg/kg q24h
Concentration (µg/mL)
Time (h)
G
Herranz - 11.95 mg/kg q12h
Concentration (µg/mL)
Time (h)
H
Herranz - 29.1 mg/kg q24h
Concentration (µg/mL)
Time (h)

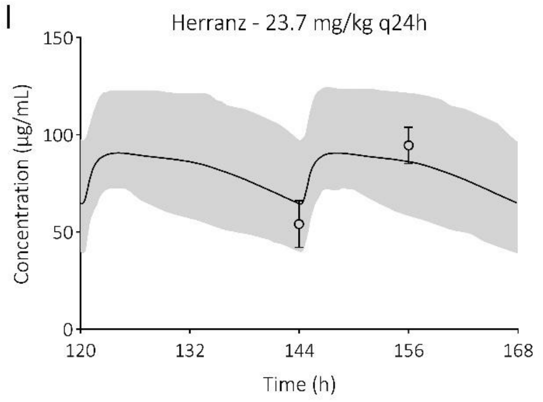

**Figure 14. Prediction of valproic acid (VPA) plasma concentration-time profiles in pediatrics upon multi-dose extended release administration.** The solid line is the predicted mean of the simulated population and the shaded area represents the $5^{th}$ to $95^{th}$ percentile of the virtual population. Open circles are the observed data: A-D (64) and E-I (65).

#### 5. Simulating alternative dosing regimens

*Methods*

After model verification, the models were applied prospectively to evaluate current dosing strategies upon oral administration in children. The current CBZ dosing regimen for epilepsy, as provided in the Dutch Paediatric Formulary (DPF), was simulated (Figure S15-17) (68). Predicted exposures are compared to the therapeutic window of 4-12 µg/ml for CBZ (69), although concerns have been raised that levels above 8 µg/ml may result in reduced cognitive function in children (70). For CBZE, the upper boundary of the reference range, i.e., 2.3 µg/ml, is included (71). To note, for CBZ in the 5-12 year simulation, simulated children weighing over 50, 40, and 33.3 kg received 1000 mg/day after 2, 3, and 4 weeks after start of the therapy, respectively. This applied to 2, 7, and 8 virtual subjects, respectively.

The current VPA dosing regimen for epilepsy, as provided in the DPF, were simulated (Figure S19-23) (72). The same dosing strategies were simulated in term neonates (Figure S21 and 23). Predicted exposures were compared to the therapeutic window of 50-100 µg/ml for VPA (69). Based on reference concentrations of 50-100 µg/ml total VPA, reference values for the free fraction are 4-15 µg/ml (i.e., 7-9% unbound at 50 µg/ml and 15% at 100 µg/ml total concentrations) (3, 73). For 4-ene-VPA, the suggested upper boundary of the reference range, i.e., 0.5 µg/ml, is included (74).

In addition, to determine the effect of altered albumin levels in children and thereby potentially necessitating different dosing strategies, the mean VPA, unbound VPA, and 4-ene-VPA concentrations were simulated under +20%, normal, -20%, and -35% altered albumin level conditions (Figure S24). The coefficient of variance (CV) % was set to zero. In addition, the (hepatic) clearances (Figure S25) and volume of distributions were plotted (Figure S26).

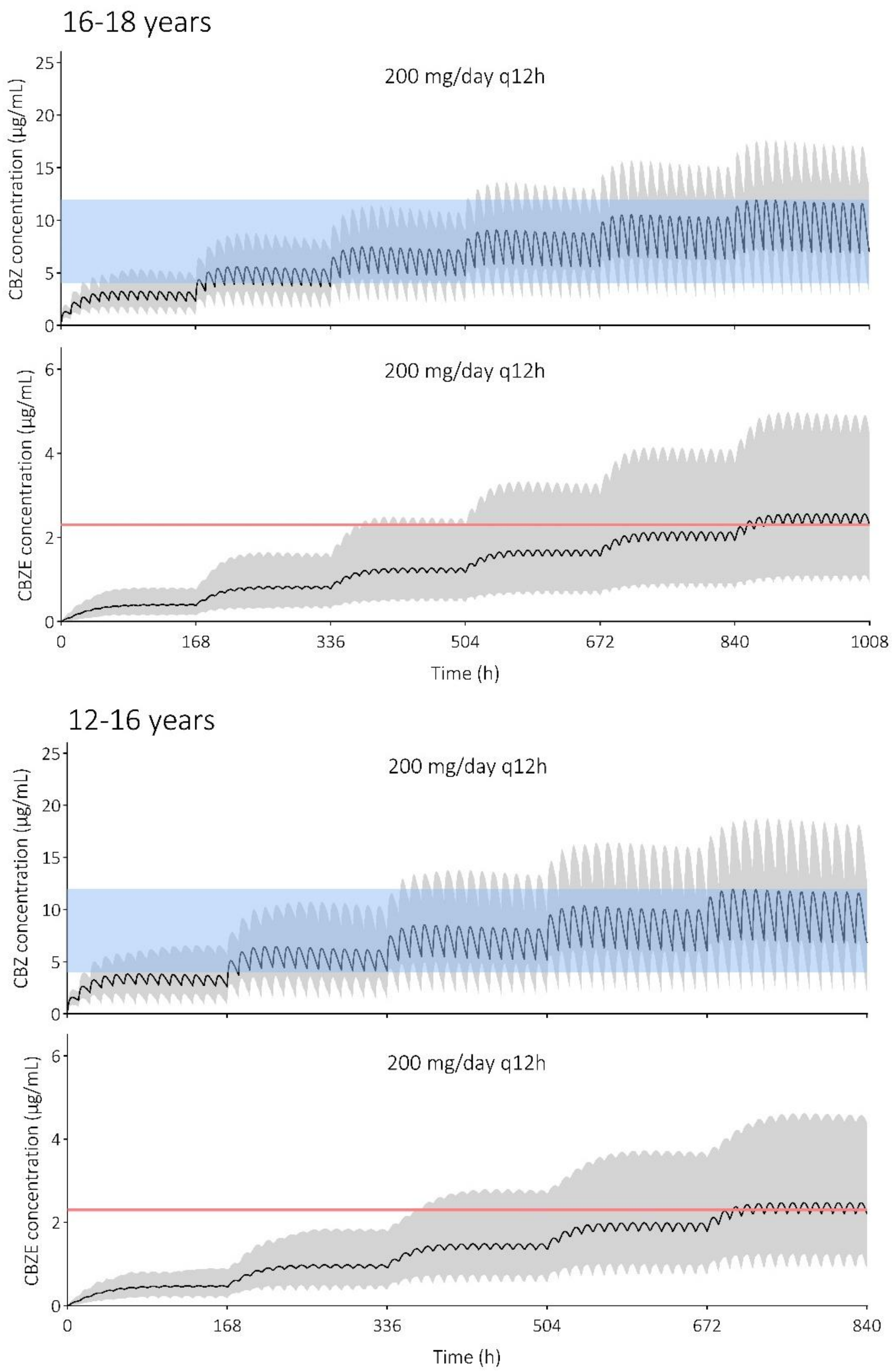

**Figure 15. Prediction of carbamazepine (CBZ) and carbamazepine-10,11-epoxide (CBZE) concentrations following the current dosing guideline in children aged 12 to 16 and 16 to 18 years.** The dose increases with 200 mg/day each week, with a maximum of 1000 mg/day and 1200 mg/day, respectively. The black solid line is the predicted mean of the simulated population and the gray shaded area represents the 5th to 95th percentile of the virtual population. The blue shaded area indicates the CBZ therapeutic window (i.e., 4-12 µg/ml) and the red solid line indicates the CBZE upper boundary of the reference range (i.e., 2.3 µg/ml).

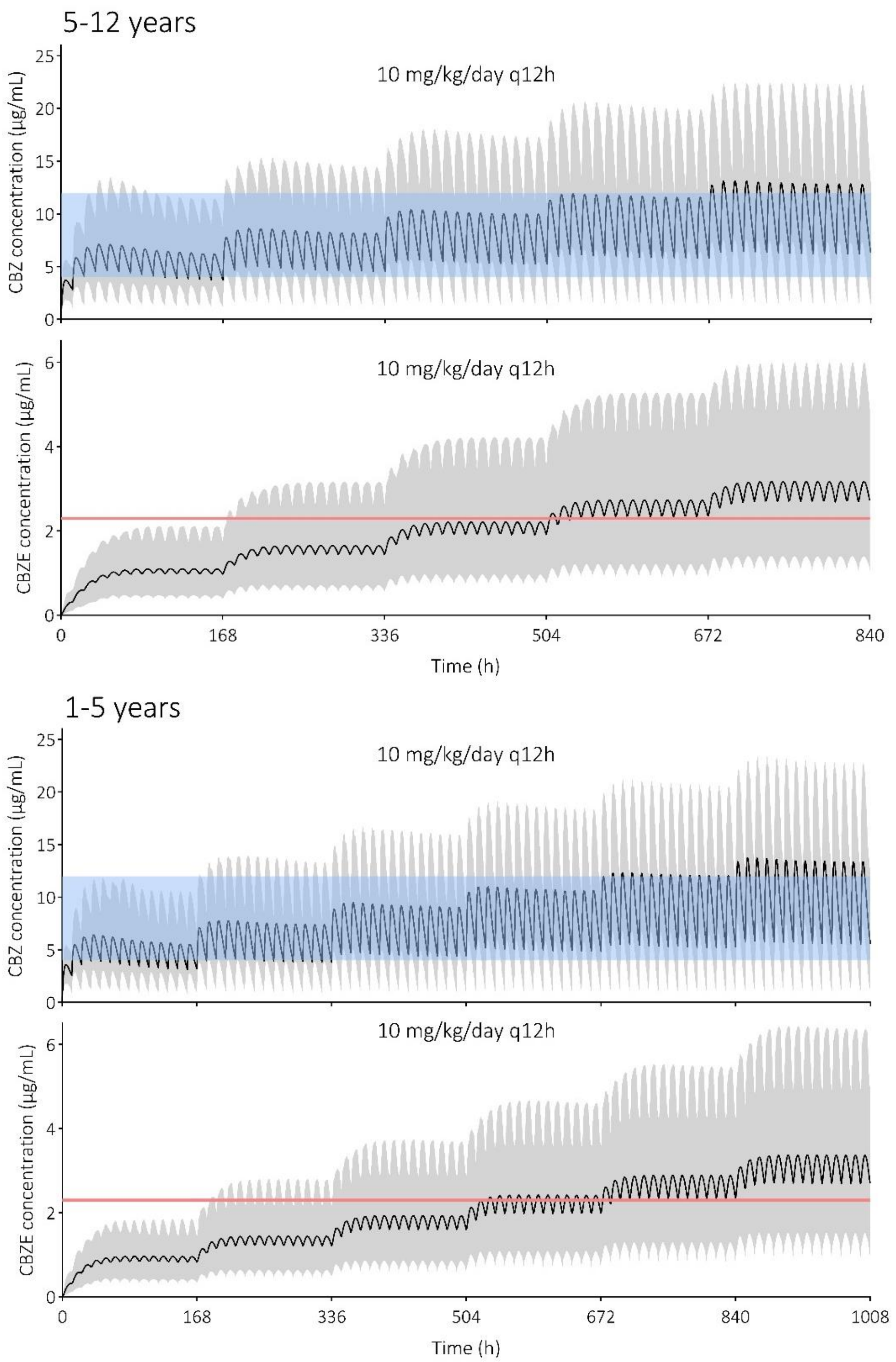

**Figure 16. Prediction of carbamazepine (CBZ) and carbamazepine-10,11-epoxide (CBZE) concentrations following the current dosing guideline in children aged 1 to 5 and 5 to 12 years.** The dose increases with 5 mg/kg/day each week, with a maximum of 35 mg/kg/day and 1000 mg/day, respectively. The black solid line is the predicted mean of the simulated population and the gray shaded area represents the 5th to 95th percentile of the virtual population. The blue shaded area indicates the CBZ therapeutic window (i.e., 4-12 µg/ml) and the red solid line indicates the CBZE upper boundary of the reference range (i.e., 2.3 µg/ml).

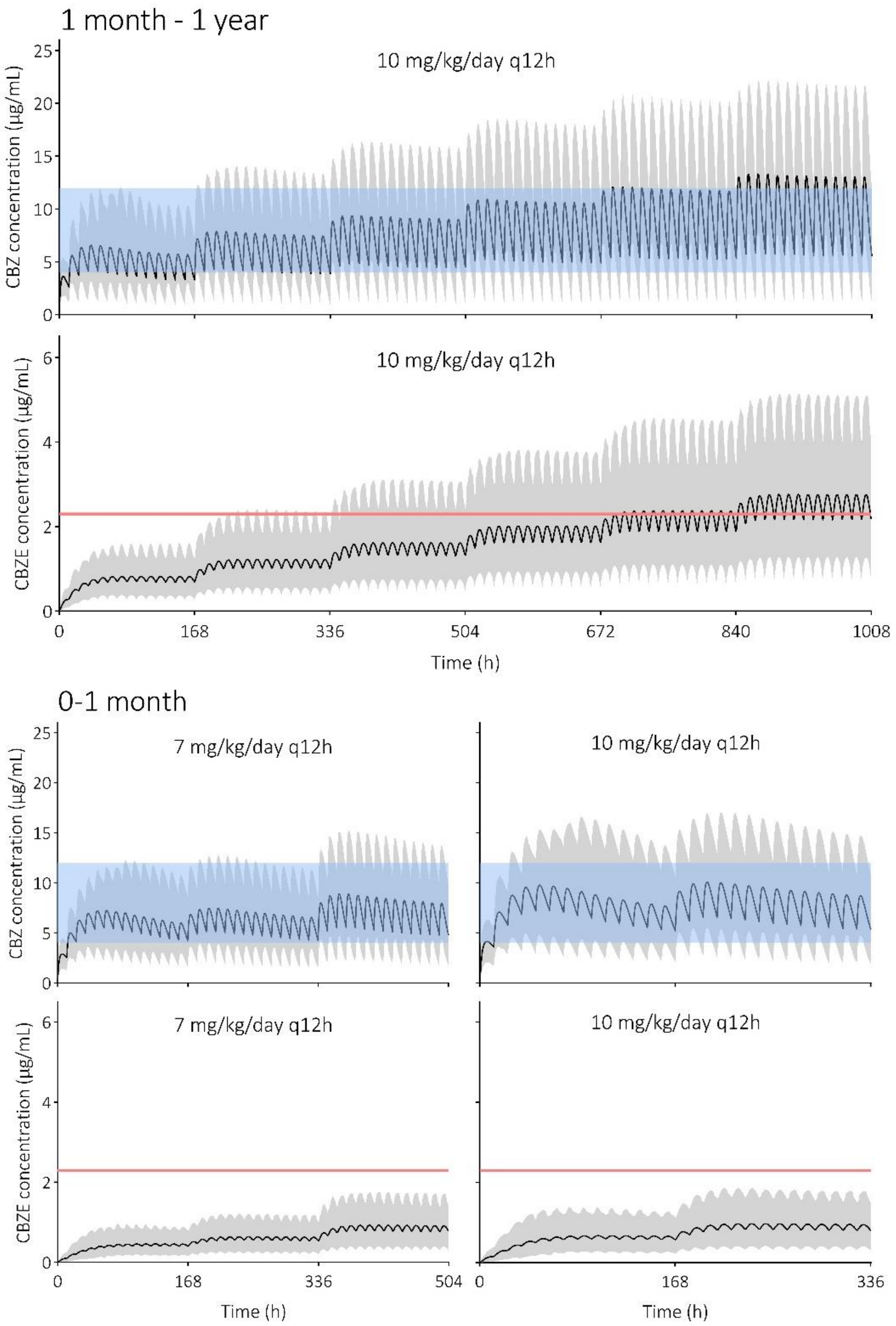

**Figure 17. Prediction of carbamazepine (CBZ) and carbamazepine-10,11-epoxide (CBZE) concentrations following the current dosing guideline in children aged 0 to 1 month and 1 month to 1 year.** The dose increases with 5 mg/kg/day each week, with a maximum of 15 mg/kg/day (where the 7 mg/kg/day dosing regimen first changes to a 10 mg/kg/day regimen) and with a maximum of 35 mg/kg/day, respectively. The black solid line is the predicted mean of the simulated population and the gray shaded area represents the 5$^{th}$ to 95$^{th}$ percentile of the virtual population. The blue shaded area indicates the CBZ therapeutic window (i.e., 4-12 µg/ml) and the red solid line indicates the CBZE upper boundary of the reference range (i.e., 2.3 µg/ml).

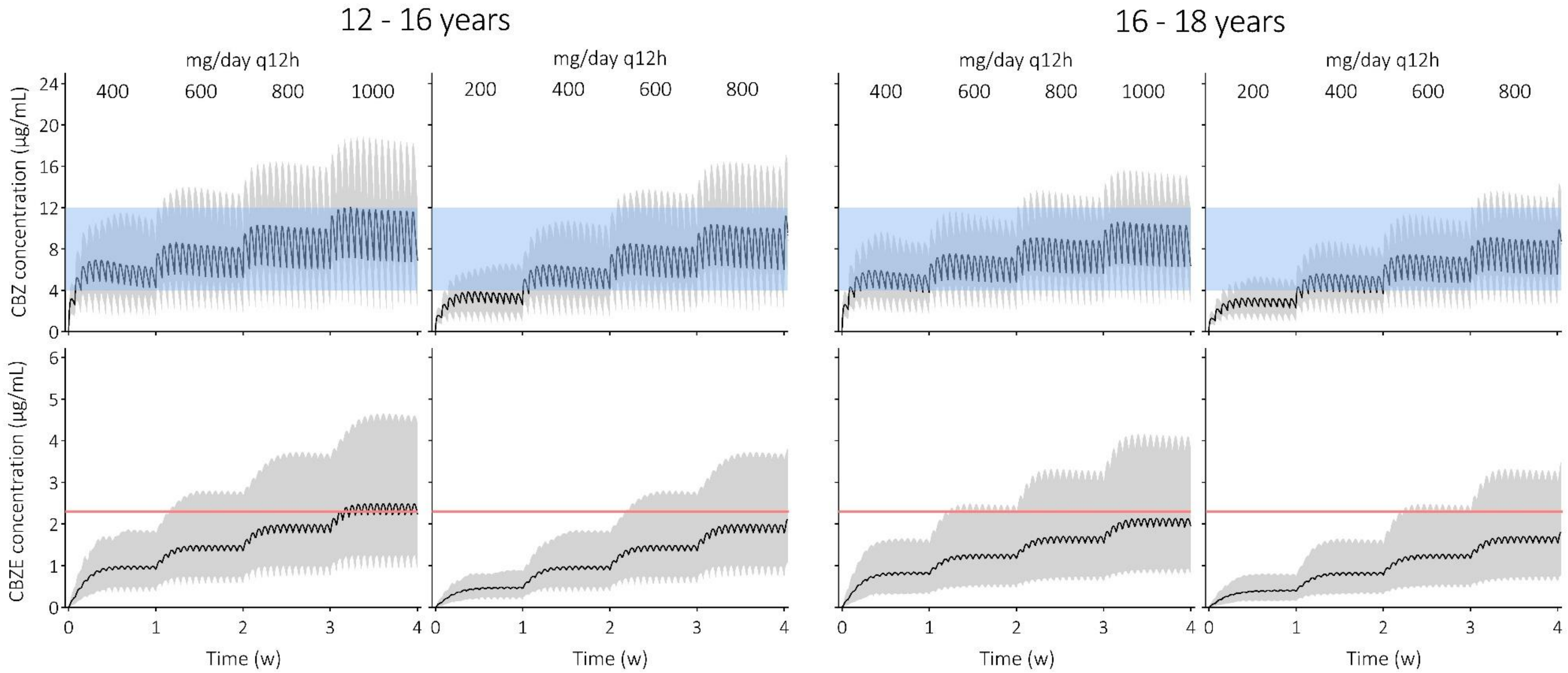

**Figure 18. Prediction of carbamazepine (CBZ) and carbamazepine-10,11-epoxide (CBZE) concentrations following the current and an adjusted dosing scheme in children aged 12 to 16 years and 16 to 18 years.** The dose increases with 200 mg/day each week. The black solid line is the predicted mean of the simulated population and the gray shaded area represents the 5$^{th}$ to 95$^{th}$ percentile of the virtual population. The blue shaded area indicates the CBZ therapeutic window (i.e., 4-12 µg/ml) and the red solid line indicates the CBZE upper boundary of the reference range (i.e., 2.3 µg/ml).

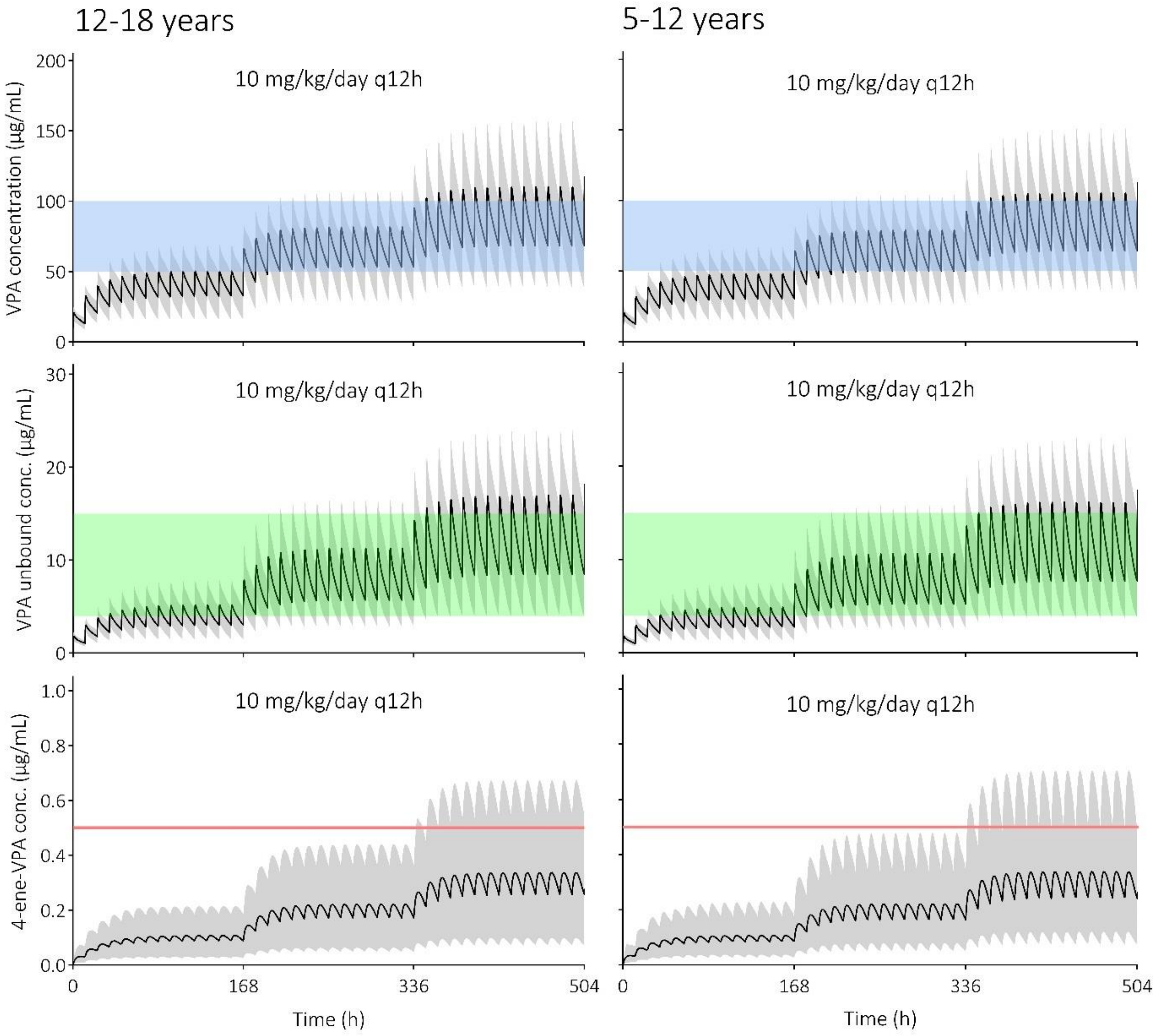

**Figure 19. Prediction of valproic acid (VPA), unbound VPA, and 4-ene-VPA concentrations following the current dosing guideline in children aged 5 to 18 years.** The dose increases with 10 mg/kg/day each week, with a maximum of 30 mg/kg/day. The black solid line is the predicted mean of the simulated population and the gray shaded area represents the 5$^{th}$ to 95$^{th}$ percentile of the virtual population. The blue shaded area indicates the VPA therapeutic window (i.e., 50-100 µg/ml), the green shaded area indicated the unbound VPA therapeutic window (i.e., 4-15 µg/ml), and the red solid line indicates the 4-ene-VPA upper boundary of the reference range (i.e., 0.5 µg/ml).

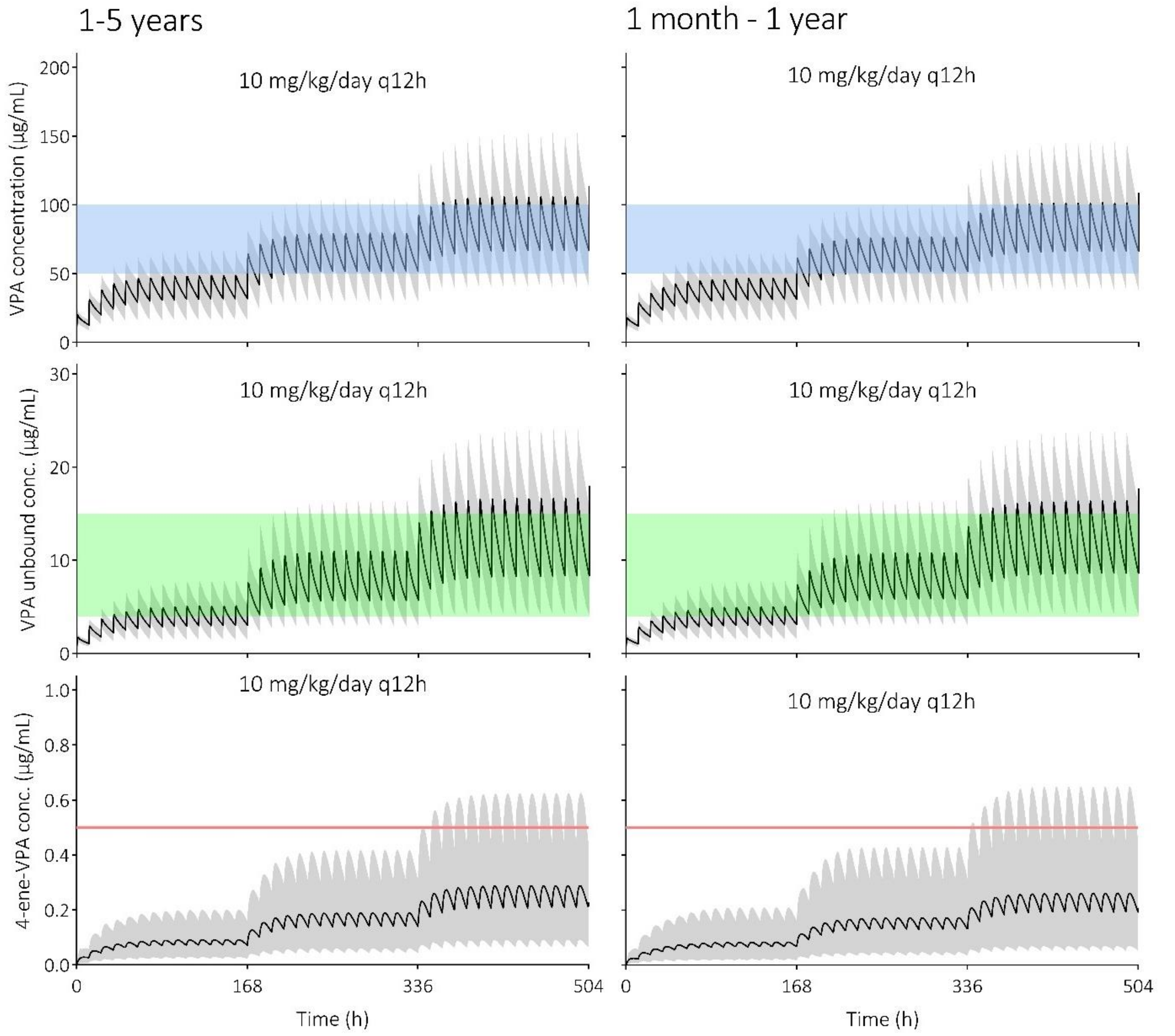

**Figure 20. Prediction of valproic acid (VPA), unbound VPA, and 4-ene-VPA concentrations following the current dosing guideline in children aged 1 month to 5 years.** The dose increases with 10 mg/kg/day each week, with a maximum of 30 mg/kg/day. The black solid line is the predicted mean of the simulated population and the gray shaded area represents the 5$^{th}$ to 95$^{th}$ percentile of the virtual population. The blue shaded area indicates the VPA therapeutic window (i.e., 50-100 µg/ml), the green shaded area indicated the unbound VPA therapeutic window (i.e., 4-15 µg/ml), and the red solid line indicates the 4-ene-VPA upper boundary of the reference range (i.e., 0.5 µg/ml).

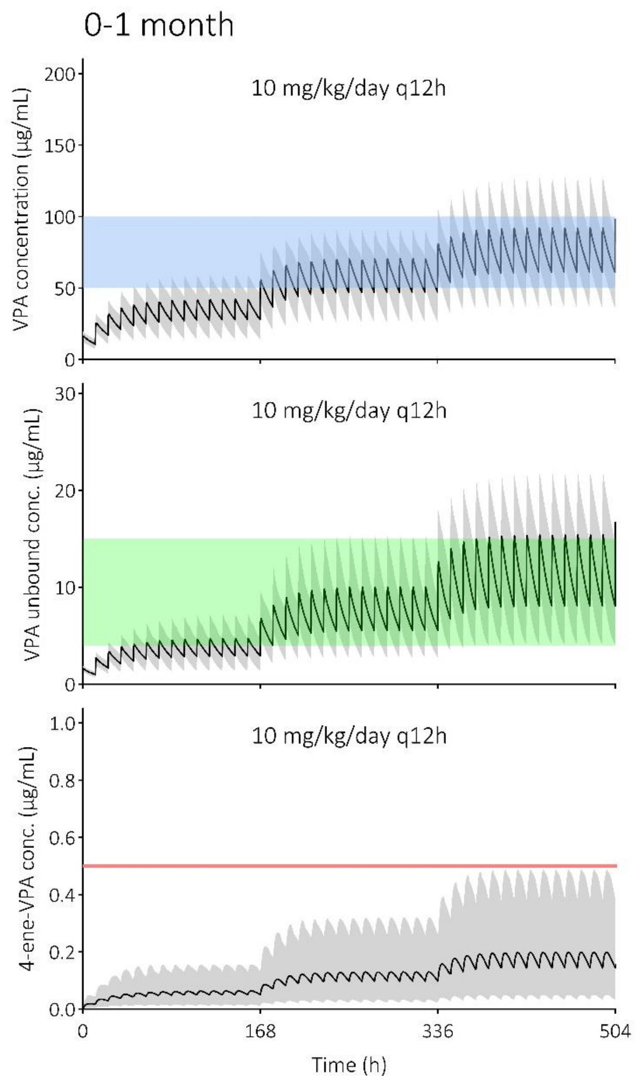

**Figure 21. Prediction of valproic acid (VPA), unbound VPA, and 4-ene-VPA concentrations following the current dosing guideline in children aged 0 to 1 month.** The dose increases with 10 mg/kg/day each week, with a maximum of 30 mg/kg/day. The black solid line is the predicted mean of the simulated population and the gray shaded area represents the 5$^{th}$ to 95$^{th}$ percentile of the virtual population. The blue shaded area indicates the VPA therapeutic window (i.e., 50-100 µg/ml), the green shaded area indicated the unbound VPA therapeutic window (i.e., 4-15 µg/ml), and the red solid line indicates the 4-ene-VPA upper boundary of the reference range (i.e., 0.5 µg/ml).

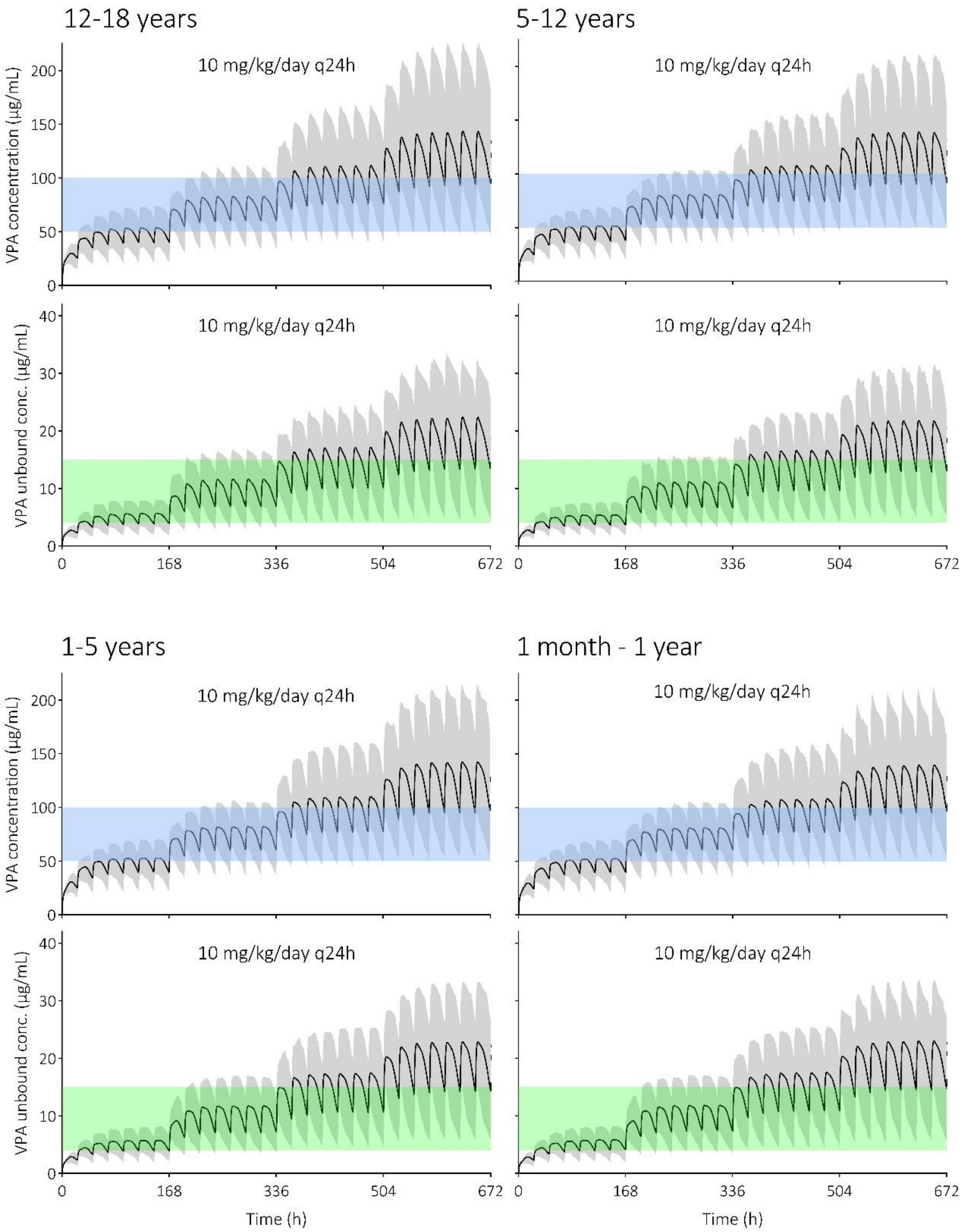

**Figure 22. Prediction of valproic acid (VPA) and unbound VPA concentrations following the current extended release dosing guideline in children aged 1 month to 18 years.** The dose increases with 10 mg/kg/day each week, with a maximum of 30 mg/kg/day. The black solid line is the predicted mean of the simulated population and the gray shaded area represents the 5$^{th}$ to 95$^{th}$ percentile of the virtual population. The blue shaded area indicates the VPA therapeutic window (i.e., 50-100 µg/ml) and the green shaded area indicated the unbound VPA therapeutic window (i.e., 4-15 µg/ml).

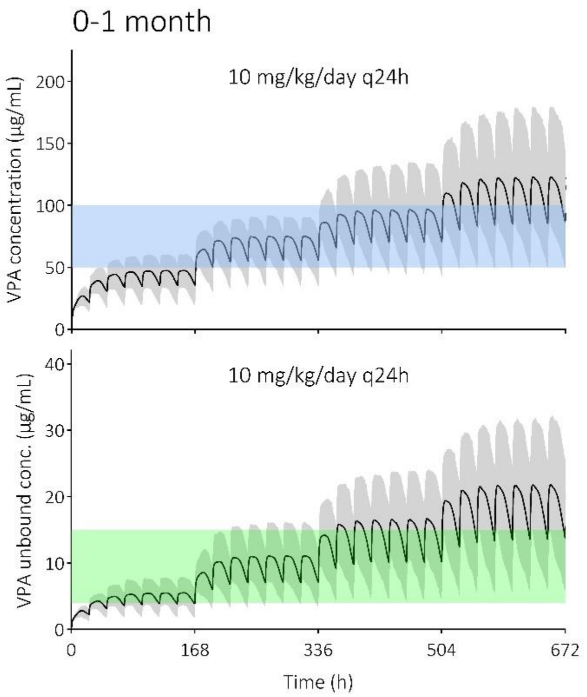

**Figure 23. Prediction of valproic acid (VPA) and unbound VPA concentrations following the current extended release dosing guideline in children aged 0 to 1 month without 'redefine subjects over time' function.** The dose increases with 10 mg/kg/day each week, with a maximum of 30 mg/kg/day. The black solid line is the predicted mean of the simulated population and the gray shaded area represents the 5th to 95th percentile of the virtual population. The blue shaded area indicates the VPA therapeutic window (i.e., 50-100 µg/ml) and the green shaded area indicated the unbound VPA therapeutic window (i.e., 4-15 µg/ml).

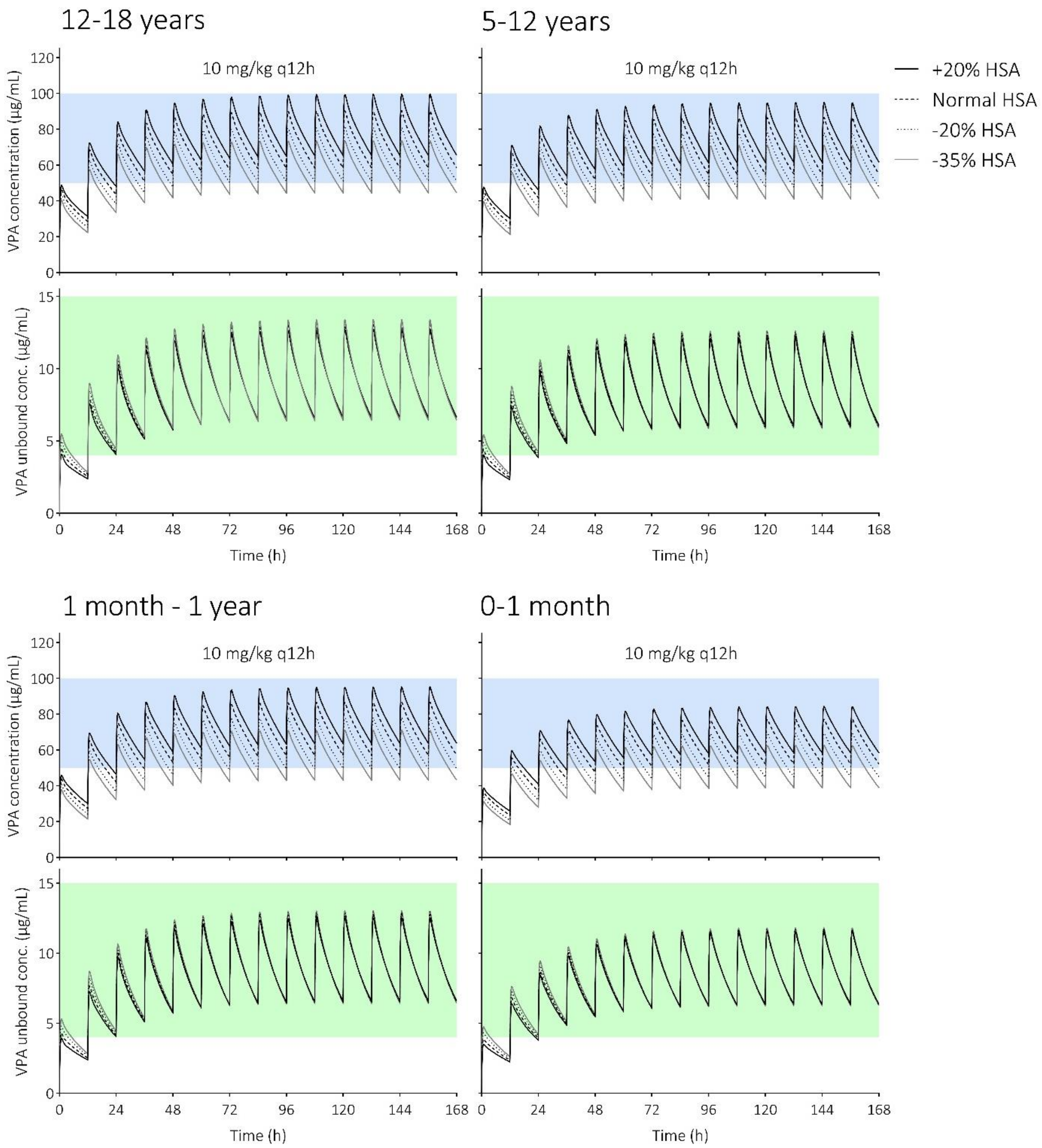

**Figure 24. Prediction of valproic acid (VPA) and unbound VPA concentrations following 10 mg/kg immediate release VPA every 12 hours (q12h) in children aged 0 to 18 years with varying human serum albumin (HSA) levels.** The lines are the predicted means of the simulated populations. The blue shaded area indicates the VPA therapeutic window (i.e., 50-100 µg/ml) and the green shaded area indicates the unbound VPA therapeutic window (i.e., 4-15 µg/ml).

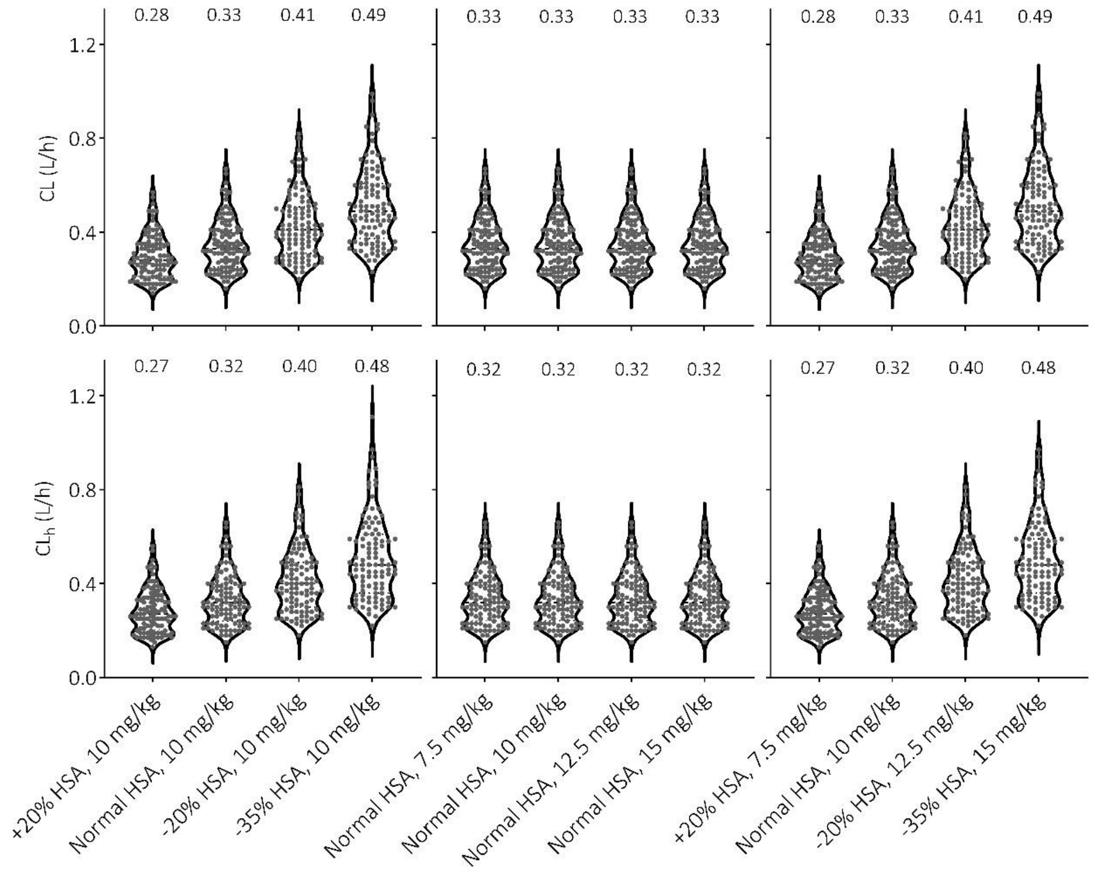

**Figure 25. Violin plots of predicted valproic acid clearance (CL) and hepatic clearance ($CL_h$) values upon either altered human serum albumin (HSA) levels, or altered doses, or both per simulated individual (n = 100, 12-18 years).** Median levels are shown on top.

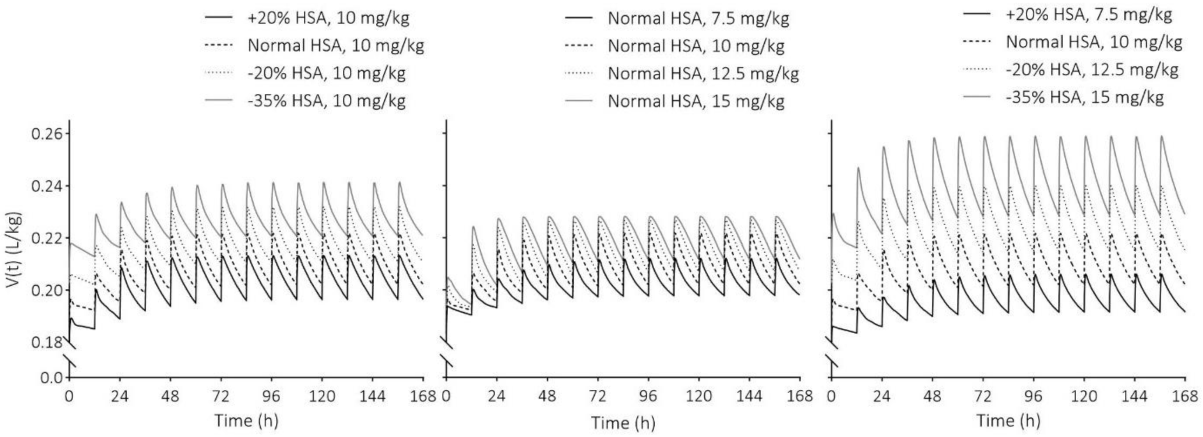

**Figure 26. Predicted volume of distributions over time upon either altered human serum albumin (HSA) levels, or altered doses, or both.** The lines are the predicted means of the simulated populations (n = 100, 12-18 years).

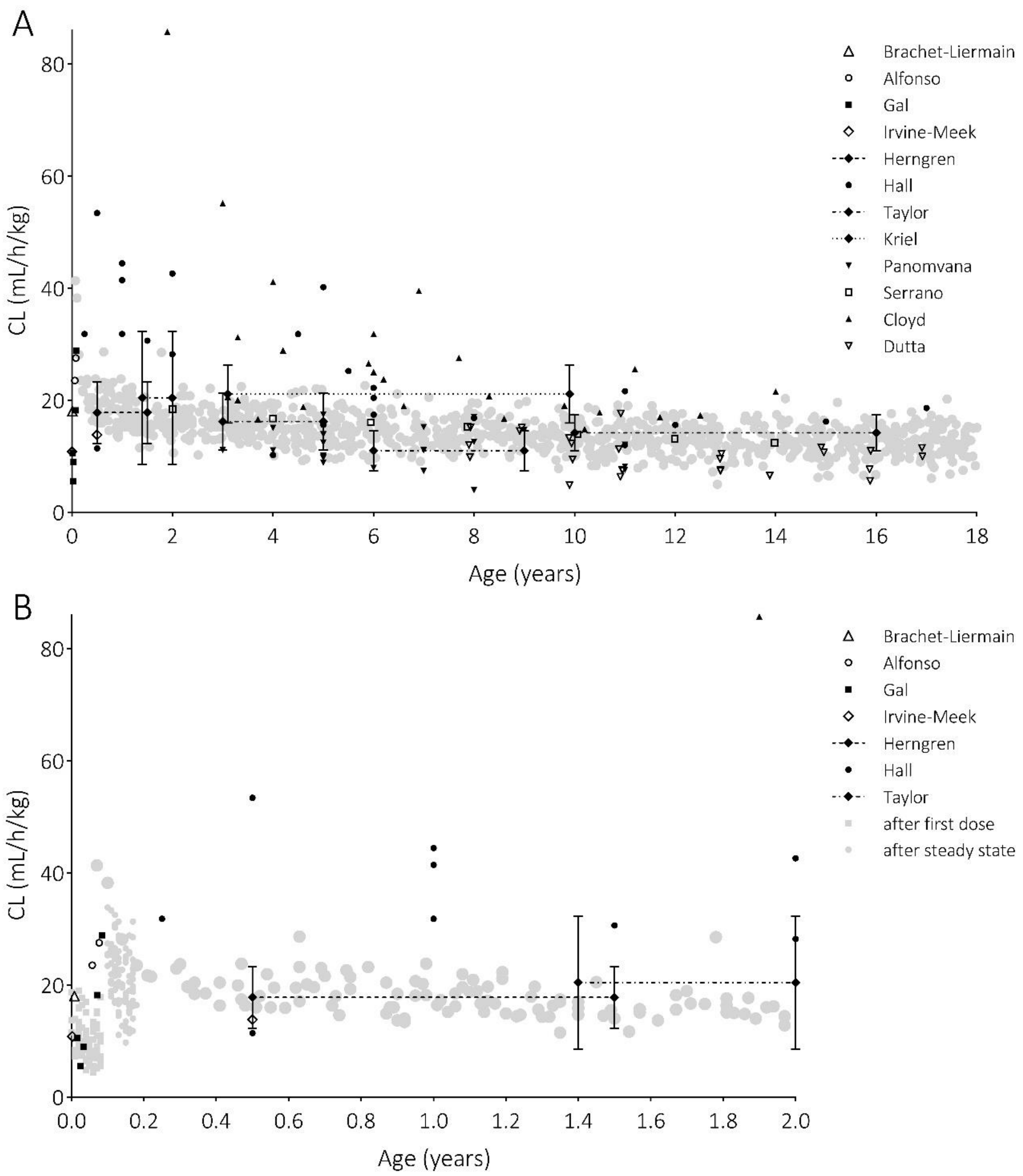

**Figure 27. Predicted and observed valproic acid clearances (CL) corrected for body weight A) across the whole pediatric age range, and B) from 0 to 2 years old.** The grey dots are individual predicted clearances after 3 weeks of treatment. For neonates, predicted clearances were provided upon a first dose and after 5 weeks of treatment. Observed data are either individual data points, or an average clearance with standard deviation across a certain age range., i.e., indicated with a connecting line. Observed data: Brachet-Liermain (51), Alfonso (46), Gal (53), Irvine-Meek (75), Herngren (54), Hall (48), Taylor (76), Kriel (77), Panomvana (78), Serrano (79), Cloyd (47), and Dutta (64).
